# The LEADING Guideline. Reporting Standards for Expert Panel, Best-Estimate Diagnosis, and Longitudinal Expert All Data (LEAD) Studies

**DOI:** 10.1101/2024.03.19.24304526

**Authors:** Veerle C Eijsbroek, Katarina Kjell, H Andrew Schwartz, Jan R Boehnke, Eiko I Fried, Daniel N Klein, Peik Gustafsson, Isabelle Augenstein, Patrick M M Bossuyt, Oscar N E Kjell

**Affiliations:** Department of Psychology, Lund University, Lund, Sweden; Department of Computer Science, Stony Brook University, New York, the United States; School of Health Sciences, University of Dundee, Dundee, Scotland; Institute of Psychology, Leiden University, Leiden, the Netherlands; Department of Psychology, Stony Brook University, New York, the United States; Faculty of Medicine, Lund University, Lund, Sweden; Department of Computer Science, University of Copenhagen, Copenhagen, Denmark; Department of Epidemiology and Data Science, Amsterdam University Medical Centers, Amsterdam, the Netherlands

**Keywords:** Expert Panel, LEAD, Best-Estimate Diagnosis, Reference standard, Criterion standard, Gold standard, Medical assessments, Psychiatric assessments, Psychological assessments

## Abstract

**Background:** Accurate assessments of symptoms and illnesses are essential for health research and clinical practice but face many challenges. The absence of a single error-free measure is currently addressed by assessment methods involving experts reviewing several sources of information to achieve a more accurate or best-estimate assessment. Three bodies of work spanning medicine, psychiatry, and psychology propose similar assessment methods: The Expert Panel, the Best-Estimate Diagnosis, and the Longitudinal Expert All Data (LEAD) method. However, the quality of such best-estimate assessments is typically very difficult to evaluate due to poor reporting of the assessment methods and when they are reported, the reporting quality varies substantially. Here, we tackle this gap by developing reporting guidelines for such best-estimate assessment studies.

**Methods:** The development of the reporting guidelines followed a four-stage approach: 1) drafting reporting standards accompanied by rationales and empirical evidence, which were further developed with a patient organization for depression, 2) incorporating expert feedback through a two-round Delphi procedure, 3) refining the guideline based on an expert consensus meeting, and 4) testing the guideline by i) having two researchers test it and ii) using it to examine the extent previously published studies report the standards. The last step also provides evidence for the need for the guideline: 10 to 63% (Mean = 33%) of the standards were not reported across thirty randomly selected studies.

**Results:** The LEADING guideline comprises 20 reporting standards related to four groups: The *Longitudinal design* (four standards); the *Appropriate data* (four standards); the *Evaluation – experts, materials, and procedures* (ten standards); and the *Validity* group (two standards).

**Conclusions:** We hope that the LEADING guideline will be useful in assisting researchers in planning, conducting, reporting, and evaluating research aiming to achieve best-estimate assessments.

## Introduction

Establishing valid and reliable assessments of symptoms and diagnoses is the foundation of health and clinical sciences. Given that reliable biological markers or specific objective signs for most mental health problems are lacking and many medical conditions only show objective markers in late stages, accurate assessments are difficult^1,2^. Essentially every single measure of a psychological construct has some potential source of bias (e.g., self-report and recall bias) or can be seen as fallible in some respect^3,4^ – which can result in inaccurate assessments and delayed treatments.

The absence of a single error-free measure can be addressed by involving multiple experts reviewing several sources of information to form a *best-estimate assessment* or a reference standard^5–7^. To understand the quality of such an assessment, it is crucial to understand how it was reached (i.e., the quality of the specific assessment method used). However, the quality of best-estimate assessments is typically very difficult to evaluate due to poor reporting of the assessment method, and when the method is reported, the reporting quality varies substantially^7^. Here we tackle this problem by developing a guideline for how to report assessment methods that aim to achieve such a best-estimate assessments, i.e., where experts review several sources of (longitudinal) information to achieve a more accurate assessment than a single, error-prone measure.

### Assessment

*Assessment* includes the evaluation, integration, and interpretation of several sources of information (e.g., outcomes of different measures, tests, or scans) to derive a valid and reliable decision (e.g., a best-estimate diagnosis)^8^. Accurate assessments are crucial for understanding the prevalence of clinical problems^9,10^, detecting and starting early treatment^11,12^, validating measurement tools^13,14^, and evaluating interventions/therapies^15,16^. In clinical practice, under- or over-estimation of illnesses can have severe negative impacts on people’s lives. In research, they threaten the validity of scientific results. For policy and implementation development, assessments are the basis for guideline development and the economic and societal evaluations of interventions. Furthermore, obtaining more accurate assessments has become increasingly important considering that high-accuracy assessments are needed in diverse fields such as Biological Psychiatry (e.g., to find reliable biomarkers linked to reference standard assessments^17–19)^ and Artificial Intelligence (e.g., to train models to reference standard assessments^20–22)^.

### A methodological solution

Here, we connect three bodies of literature that have proposed similar assessment methods: The Expert Panel method in medicine^7,23,24^ – as well as the Best-Estimate Diagnosis^6,25^ and the Longitudinal Expert All Data (LEAD)^5^ methods in psychiatry and clinical psychology. The three methods share the same goal of attaining best-estimate assessments through similar methodological approaches: All three methods use expert panels or consensus teams (e.g., clinical psychologists or medical doctors) to review several sources of information (e.g., clinical questionnaires and medical tests) to establish a more accurate assessment (e.g., for diagnostic purposes in clinical practice, or as a reference standard in statistical modeling). The three methods accentuate different parts of this methodological approach: The Best-Estimate Diagnosis method accentuates the use of informants and objective tests next to self-reported data^6,25^; the Expert Panel method focuses on the characteristics, constitution, and procedure of the panel^7,23^ and only the LEAD method requires a longitudinal design^5^, although longitudinal data are also used in some Expert Panel designs (≈27% of studies^7^). Herein we collectively refer to these three approaches as the *assessment methods*.

The result of the *assessment methods* is a consensually derived criterion (e.g., a best-estimate assessment) that has been used for many different applications where there is no single error-free measure. It has, for example, been used to i) understand the accuracy of a measurement tool or marker through comparison to a best-estimate assessment^26–31;^ ii) establish the prevalence of symptoms and disorders^9,10,32^; iii) establish the temporal stability or development of symptoms and disorders ^33–35^; iv) improve (earlier) detection or screening of symptoms or disorders^11,12,36^; v) study genetics and family history^37–39^; and vi) examine classification systems or diagnostic criteria^40–42^. The applications span diverse fields, including medicine, psychiatry, clinical psychology, public health/epidemiology, and artificial intelligence. Box 1 provides more examples of how the assessment methods have been applied in different types of studies across fields.

##### Box 1. Overview of applications of the assessment methods across different study designs and fields

The absence of a single error-free measure can be mitigated by involving multiple experts reviewing several sources of information to form a best-estimate assessment. To assist users in planning, evaluating, and reporting assessment method procedures to derive such best-estimate assessments, we developed a guideline with stakeholders from areas in which the assessment methods are used, such as medicine, psychiatry, clinical psychology, and epidemiology. The use cases below show study designs and fields where the guideline is applicable and relevant for reporting the assessment method procedure. For example, the assessment methods have been applied:

**1. To evaluate a measure’s accuracy against a reference standard.** To understand the accuracy of a measurement tool, there is a need to compare it to a more accurate or best-estimate assessment. For example, it has been used:

- *in psychiatry*, for evaluating MINI-KID diagnoses for children and adolescents^26^

*and* evaluating DSM diagnoses in patients with psychosis.^27^

- *in clinical psychology*, for evaluating Major Depression Inventory (MDI) severity scores.^28^

- *in medicine*, for evaluating deep learning models assessing liver cancer^29^

*and* evaluating prediction rules for coronary artery disease.^30^

- *in public health/epidemiology*, for evaluating electronic health record algorithms for assessing asthma.^31^

**2. To establish the prevalence of symptoms or disorders.** For example, it has been used:

- *in public health/epidemiology*, for assessing the prevalence and familiality of pathological gambling.^9^

- *in psychiatry*, for assessing the prevalence of eating disorders in patients with personality disorders.^10^

- *in medicine*, for assessing the prevalence of clinically relevant incidental findings when diagnosing pulmonary embolism.^32^

**3. To establish the temporal stability or development of symptoms or disorders.** For example, it has been used:

- *in clinical psychology*, for learning about autism spectrum disorder diagnoses during childhood^33^

*and* for learning about the course of bipolar disorder.^34^

- *in psychiatry*, for assessing diagnostic stability in individuals with autism spectrum disorder.^35^

**4. To improve (earlier) detection or screening of symptoms or disorders**. For example, it has been used:

- *in psychiatry*, for the assessment of personality disorders.^11^

- *in medicine*, for the early detection of heart failure.^36^

*and* for early detection of injuries in physically abused older adults.^12^

**5. To study genetic history and family heritability.** For example, it has been used:

- *in psychiatry*, for learning about genetic risks for ADHD.^37^

- *in clinical psychology*, for studying familial aggregation and heritability of subtypes of depression^38^

*and* for studying the familial transmission of mania and depression.^39^

**6. To examine classification systems or diagnostic criteria.** For example, it has been used:

- *in clinical psychology*, for evaluating the DSM criteria for hoarding disorder.^40^

- *in psychiatry*, for examining a DSM alternative model for personality disorders^41^

*and* for comparing DSM-IV and -5 criteria of autism spectrum disorders.^42^

**Description of an example study including a best-estimate assessment**

Best-estimate assessments have, for example, been established to evaluate the validity of the Schedule for Affective Disorders and Schizophrenia for School-Age Children-Present and Lifetime version (K-SADS-PL; a semi-structured diagnostic interview used in child and adolescent psychiatry)^43^. The best-estimate assessments were diagnoses of neurodevelopmental and related disorders made by five experienced child psychiatrists based on the DSM-5 criteria. To achieve best-estimate assessments, patients were followed for at least three months, and the psychiatrists had access to all available data (except for the K-SADS-PL diagnoses), including information from medical records, interviews, questionnaires, laboratory tests, as well as information provided by clinical staff, caregivers and teachers. Criterion validity of the K-SADS-PL was established as the agreement of the diagnoses with the best-estimate assessments.

### Reporting issues

The assessment methods possess a high potential for achieving best-estimate reference standards in many situations. However, the quality of such proclaimed best-estimate assessments varies substantially and is typically very difficult to evaluate due to poor reporting of the method *how* they were achieved (e.g., see reviews of expert panels^7,23^). A systematic review of assessment methods and reporting of expert panels^7^ has demonstrated that the methods used for panel or consensus diagnoses vary substantially across studies and that many aspects of the procedure are often unclear or not reported at all. Many recent studies fail to report central aspects of the three assessment methods, including the quality, structure, or presentation of the data^44^, the training and qualifications of the experts^45^, the method for avoiding biases and achieving consensus^13^, and the time span of the longitudinal design-component^46^. The poor operationalization of the assessment methods jeopardizes the goal of achieving best-estimate assessments – where a vaguely described method makes it difficult to evaluate the research. Referring to an assessment as a best-estimate (and sometimes even as a gold standard) while vaguely describing or poorly operationalizing the method for achieving the assessment is alarming^47,48^.

### The degree of validity

These assessment methods aim to achieve high validity (i.e., the degree to which the assessment captures what it aims to measure). Typically, the assessment methods aim to achieve as high validity as possible (i.e., a “leading” assessment), or, depending on resources, at least more accurate than a single error-prone measure. Despite this central aim, research often fails to clearly describe the degree of validity of the attained assessment. Using these assessment methods does not automatically guarantee high validity – it depends on how well the method is executed.

In addition, the assessments are often described with different terms: *reference standard* is often used in medicine, and *criterion standard* or *best-estimate diagnosis* is often used in psychology. We propose that the reporting of these assessment methods benefit from more explicitly describing *what* was measured and *how* well it measures up to different standards – whether and how they relate to a state-of-the-art assessment. Whereas *reference* and *criterion standards* fail to convey an intention of “nearing” a state-of-the-art assessment, the *best-estimate diagnosis* narrowly focuses on the classification of a diagnosis and not on symptom severity. Therefore, we here use the term *best-estimate assessment* in the context of describing a “leading”, state-of-the-art assessment.

### Reporting standards

Previous well-established guidelines have focused on the complete reporting of specific study designs, such as the *STrengthening the Reporting of OBservational studies in Epidemiology* (STROBE)^49^ for observational studies; the *Statement for Reporting for Diagnostic Accuracy* (STARD)^50^ for diagnostic accuracy studies; the *Consolidated Standards of Reporting Trials* (CONSORT)^51^ for randomized trials, and the *Transparent Reporting of a multivariable prediction model for Individual Prognosis Or Diagnosis* (TRIPOD)^52^ for prediction model studies (see the supplementary material [SM] for other relevant guidelines). The STARD guidance is most closely related to the reporting of the assessment methods since best-estimate assessments are often used to evaluate a measure’s (diagnostic) accuracy. However, none of the guidelines are sufficient for complete reporting of the assessment methods, where (multiple) experts review several sources of (longitudinal) information to form a best-estimate assessment. Although an earlier systematic review identified and structured the various choices involved in Expert Panel procedures^7^, no attempt was made to develop a formal guideline for the reporting of Expert Panel assessments.

### Aim

Our aim is to develop reporting standards for comprehensive reporting of these assessment methods – which can help researchers plan, carry out, and report studies employing these assessment methods, as well as help readers evaluate them. We call the reporting guideline the LEADING guideline, emphasizing the methodological components and the importance of describing *what* was assessed and *how well* (i.e., how it relates to a “leading” assessment). We further revise the original meanings of LEAD (*Longitudinal, Expert, All Data*^5^) to *Longitudinal*, *Evaluation – experts, materials and procedures*, and *Appropriate* Data). In short, the LEADING guideline aims to guide the reporting of assessment methods to improve evaluations of the assessment standard.

## Methods

### Development stages

We developed the reporting guideline over four stages: 1) drafting reporting standards; 2) incorporating expert feedback; 3) refining the final guideline, and 4) testing the guideline. The development method largely followed Moher and colleagues’ guidance for developing reporting guidelines^53^ (See Table S1 for elaborations on each recommended step). For organizational purposes, a working group (V.E., K.K., & O.K.) was set up, and a steering group (H.A.S., J.B., E.F., D.K., P.G., I.A., & P.B.) was formed to provide a wide range of expertise. The steering group included seven experts and was selected to cover a diverse range of expertise and fields related to the assessment methods (e.g., psychiatry/clinical psychology, medicine, epidemiology/public health, and Artificial Intelligence). See the declarations section for information regarding ethics.

### Drafting reporting standards

The working group drafted the original reporting standards. First, the working group, with the support of the steering group members, identified relevant research using or describing the assessment methods, including the three bodies of literature: Expert Panel^7^, Best-Estimate Diagnosis^6^, and LEAD^5^. Second, articles using any of the three assessment methods were identified through a literature search using Google Scholar with the following search terms: [“expert panel diagnosis” OR “expert panel assessment” OR “expert panel consensus” OR “expert panel methodology” OR “expert panel standard” OR “expert panel reference”] for Expert Panel studies; [“best-estimate diagnosis” OR “best-estimate diagnostic” OR “best-estimate standard” OR “best-estimate assessment” OR “best-estimate methodology” OR “best-estimate reference”] for Best-Estimate Diagnosis studies; and [“longitudinal expert all data” OR “longitudinal evaluation all data”] for LEAD studies. Articles that clearly stated the use of one of the three assessment methods were selected. Articles stating another purpose than assessment (e.g. when an expert panel was used to reach a consensus about a treatment strategy) were excluded.

Third, relevant reporting guidelines and systematic reviews were identified, including a review of expert panels applications^7^; the STROBE statement^49^; and the STARD guidance^50^ (other complementary reporting guidelines and systematic reviews are presented in the SM). The aim was for the reporting standards in the LEADING guideline to complement rather than repeat these guidelines (i.e., new standards should extend or complement existing standards rather than repeat them^53^). The use of multiple reporting guidelines may often be appropriate; for example, when reporting a randomized trial that includes best-estimate assessments, one may use CONSORT^51^ to report the trial design and main results, the LEADING guideline for describing the specifics for reaching the best-estimate assessment, and the *Consolidated Health Economic Evaluation Reporting Standards* (CHEERS)^54^ for reporting the economic evaluations and comparisons.

Potential standards were drafted by the working group with the objective of encompassing a comprehensive reporting of the assessment methods. The reporting standards were grouped into four groups: *Longitudinal design*, *Appropriate data*, *Evaluation – experts, materials and procedures*, and *Validity*. Empirical and theoretical inclusion rationales were stated for the groups and the individual standards (i.e., explanations and elaborations). Lastly, the standards with inclusion rationales were further developed through a workshop with a patient organization for depression, followed by receiving feedback from the steering group members to receive a wide range of perspectives early in the process.

### Incorporating expert feedback

To systematically collect expert feedback from different perspectives, we used a consensus-building procedure called the *Delphi technique*^55^. We used an iterative process based on two rounds of questionnaires (i.e., Delphi surveys), enabling feedback from round 1 to feed into round 2. Delphi participants received relevant background research, the reporting guideline aims, as well as the groups and the individual standards with their inclusion rationales. They provided feedback through open- and closed-ended response formats. Through open-ended responses in Round 1, experts could propose new standards and provide feedback on the formulations of existing standards, their inclusion rationales, and evidential support. In addition, two closed-ended questions^56^ about standard inclusion (*This item should be included in the reporting checklist*) and perception of study quality (*Whether this information is present or not would influence my perceptions of the quality of a study*) were answered with rating scales ranging from 1 = *Strongly disagree* to 7 = *Strongly agree*. In Round 2, the experts were asked to rate the updated reporting standards using the same two closed-ended questions as in Round 1 and to provide feedback through open-ended responses (e.g., feedback on the clarifications and reformations of the standards that resulted from Round 1).

The first and/or last authors of articles since 2013 using any of the three assessment methods were identified using the search terms described above (*n* = 87 articles, *n* = 124 authors; see the SM for more details). These authors and the seven steering group members were invited via email to participate in the Delphi Round 1 (*n* = 131 participants emailed). In total, 27 participants completed the survey (response rate 21%). Only participants from Round 1 who provided their contact details were invited to Round 2 (*n* = 25). In total, 20 participants completed the survey (response rate 80%). All participants provided their informed consent. Figure 1 presents the research experiences and demographics of the Delphi participants. Participants reported a wide range of academic backgrounds (e.g., Medicine, Psychiatry or Clinical Psychology, Artificial Intelligence, Journal Editors), and an extensive variety of relevant methodological experiences (e.g., Biological Markers, Ecological Momentary Assessments, and Expert Panels; Figure 1), with an age range of 30 – 70 years (*M* = 51.54, *SD* = 12.40).

**Figure 1.**
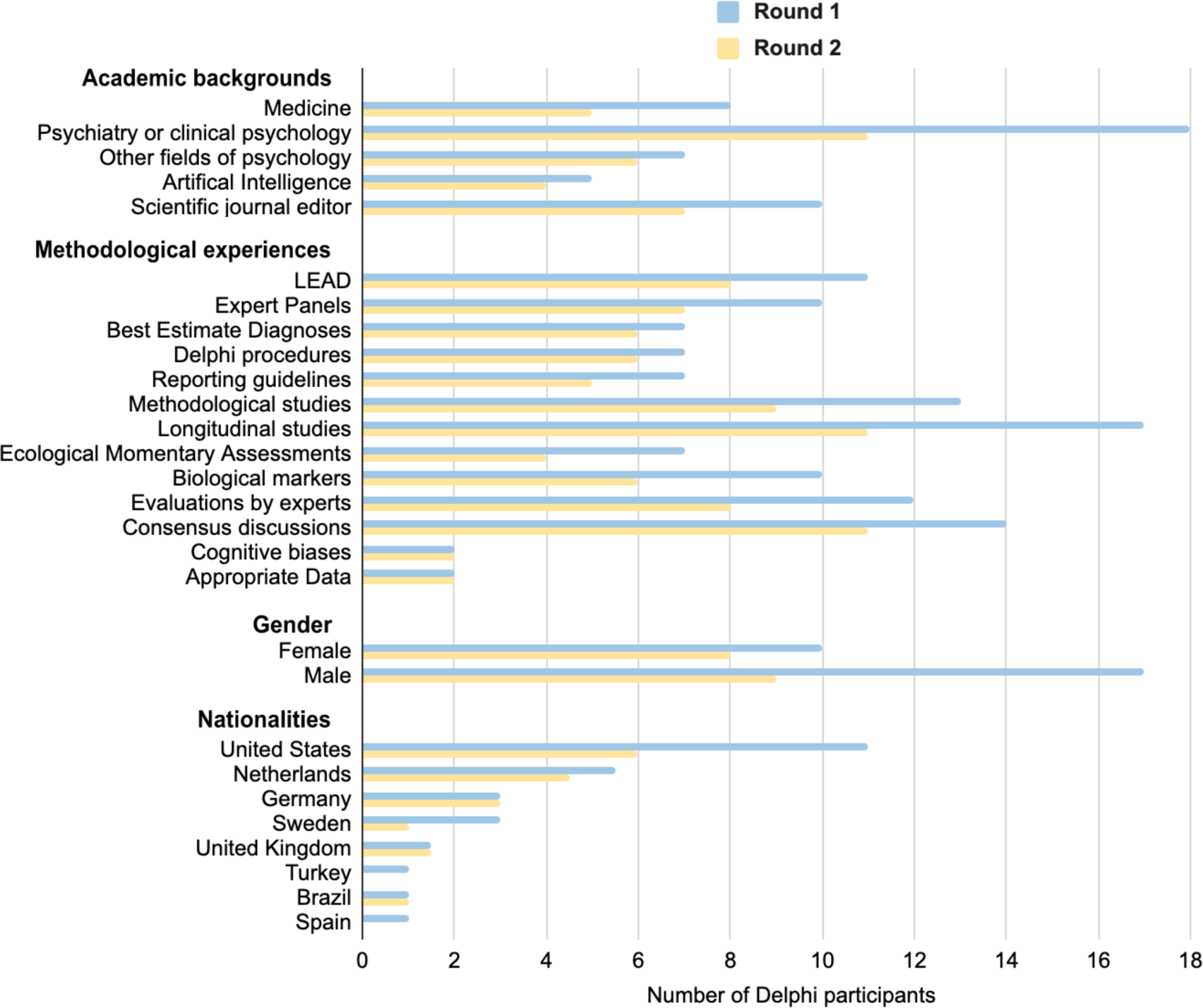
**Research experiences and demographics of the Delphi participants.** Notes. The answer options for Academic backgrounds and Methodological experiences were not mutually exclusive (i.e., multiple backgrounds and experiences could be reported by the participants). In Round 2, the demographics and reported experiences are known for 17 of the 20 participants.

*Delphi survey results.* The criteria for including a reporting standard was that the median of Delphi expert responses was at least *6 = Agree* on the question about its inclusion. In Round 1, the mean ratings for the *item inclusion* scale ranged from 5.37 - 6.67 (*M* = 6.06; *SD* = 0.31; Table S2) with a median agreement ranging from *6 = Agree* to *7 = Strongly Agree.* No new standards were suggested. The feedback resulted in the removal of one reporting standard and the clarification and reformulation of 20 standards. The standard *on Transparency and replicability* was rated as relevant but removed because it is achieved by reporting the other reporting standards. Standard *4.2 Validity and Standard* needed a major clarification about the meaning of validity as well as standard. Minor clarifications and reformations, such as grammar, or word changes, were made for 19 standards (see open material). The mean ratings in Round 2 ranged from 5.47 – 6.70 (*M* = 6.20; *SD* = 0.37; Table S3), with open feedback resulting in minor clarifications and reformulations of nine standards.

### Refining the final guideline

The authors finalized the guidelines in an expert consensus meeting. The meeting was held online with nine working and steering group members. The content and structure of the consensus meeting were prepared by the working group, and the meeting was led by the last author (O.K.). Participants had access to the guidelines, elaboration and explanation (inclusion) rationales, and the drafted paper before the meeting, where they also had the option to provide comments and feedback in writing. The meeting included going through the Delphi Rounds 1 and 2 findings and discussing the paper draft, including the individual reporting standards and groups. We decided not to carry out another Delphi round since i) the median agreement for each reporting standard in both Delphi Rounds 1 and 2 ranged from *6 = Agree* to *7 = Strongly Agree*, ii) no new standards were suggested, and iii) only minor changes were needed after Round 2, which taken together suggest consensus.

### Testing the guideline

To pilot the applicability of the guideline, the guideline was tested i) by independent researchers with experience of each method piloting the reporting of each standard and ii) by the authors (V.E., K.K.) using it to evaluate published articles. The two test procedures resulted in adding minor clarifications to three standards (*2.4 The access to the index measure, 3.3 Blindness and conflict of interest,* and *3.4 Instructions and training*). Also, a concrete example of how to report the items was added to the general guideline instructions.

*Incorporating test-user feedback.* Two test users (Ph.D., with experience using the LEAD and Expert Panel method) who had not been involved in the development of the guideline (e.g., in the Delphi procedure) were recruited to test the guideline (see the SM for more details). They were asked to report each standard based on a finished, ongoing or planned study using one of the asssessment methods and/or provide feedback about the formulation of the standards.

*Reports of the standards in 2022 and 2023.* Three separate targeted searches (LEAD, Expert-panel, Best-estimate) were conducted using the search terms described above, and the first author (V.E.) examined which standards were reported in 30 randomly selected articles applying the assessment methods in 2022 and 2023 (i.e., five from each method from each year; see the SM for the selection process). Each reporting standard was rated using four categories: standard *not reported*; standard *reported vaguely or insufficiently*; standard *(minimally) sufficiently reported*; or standard *not applicable to the study*. Out of the 30 articles, six were randomly selected (i.e., one from each method from each year) and examined by the second author (K.K.) to get insight into the accuracy of the ratings of the first author. Discussing their disagreements to reach consensus resulted in changing 23 ratings (19%) of the first author. This testing procedure also provided information about the strengths and shortcomings of contemporary reporting of published articles using these methods (see Results section).

## Results

The reporting guideline is presented in Table 1 (see Figure 2 for an overview). It comprises 20 standards for comprehensive reporting of the assessment methods divided into four groups: 1. *The Longitudinal design* group (4 standards), 2. *The Appropriate data* group (4 standards), 3. *The Evaluation – experts, materials, and procedures* group (10 standards), and 4. *The Validity* group (2 standards). The reporting standards encourage researchers to elaborate on what was done and why – whilst avoiding normative standards, such as a minimum number of experts. Each standard description in Table 1 is accompanied by an example. Further *Explanations and Elaborations* regarding the individual reporting standards and the four groups are presented in the SM, including Tables S4 and S5.

**Figure 2.**
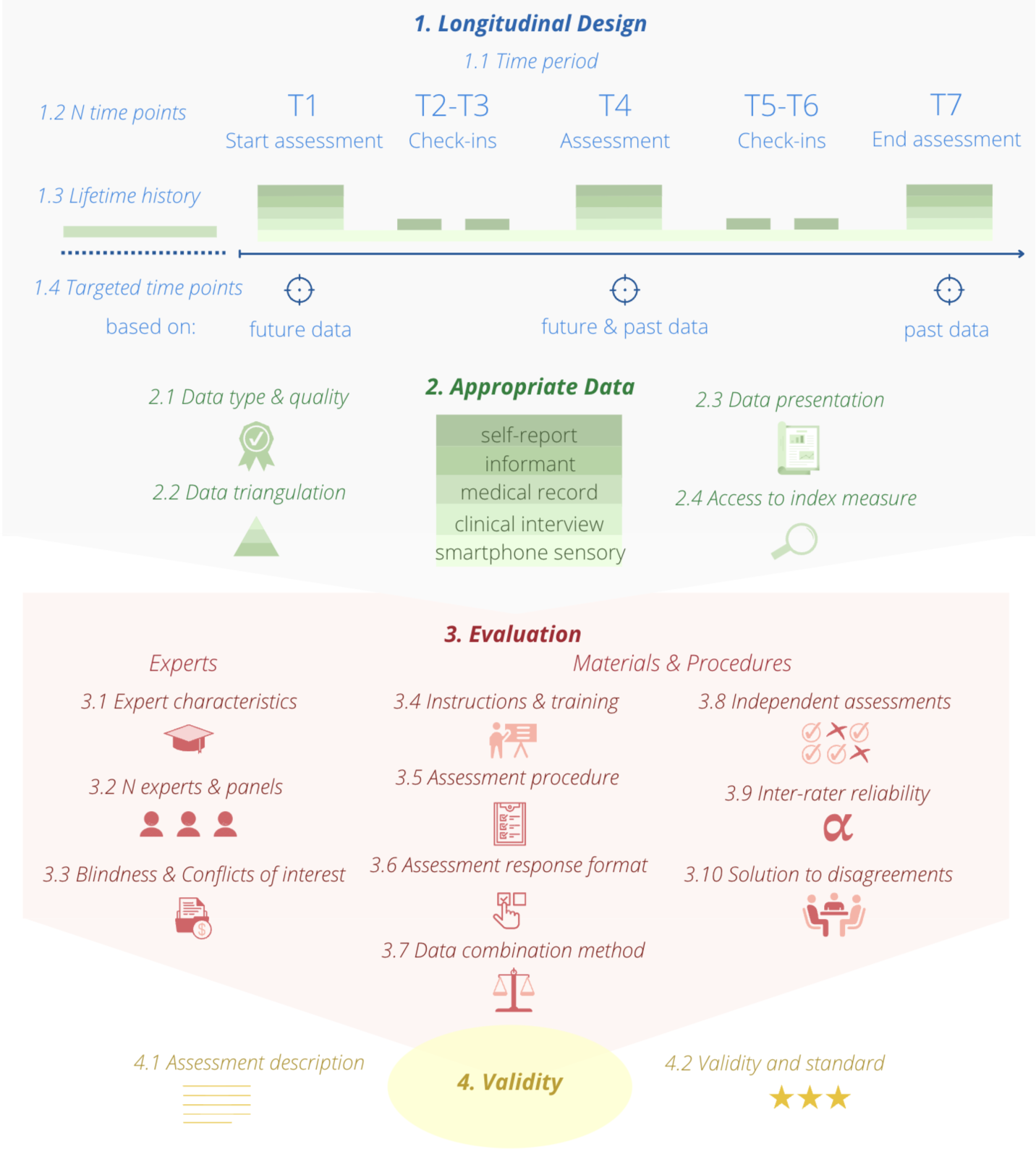
Overview of the LEADING guideline reporting standards. For more details about each standard, see Table 1.

**Table 1.**
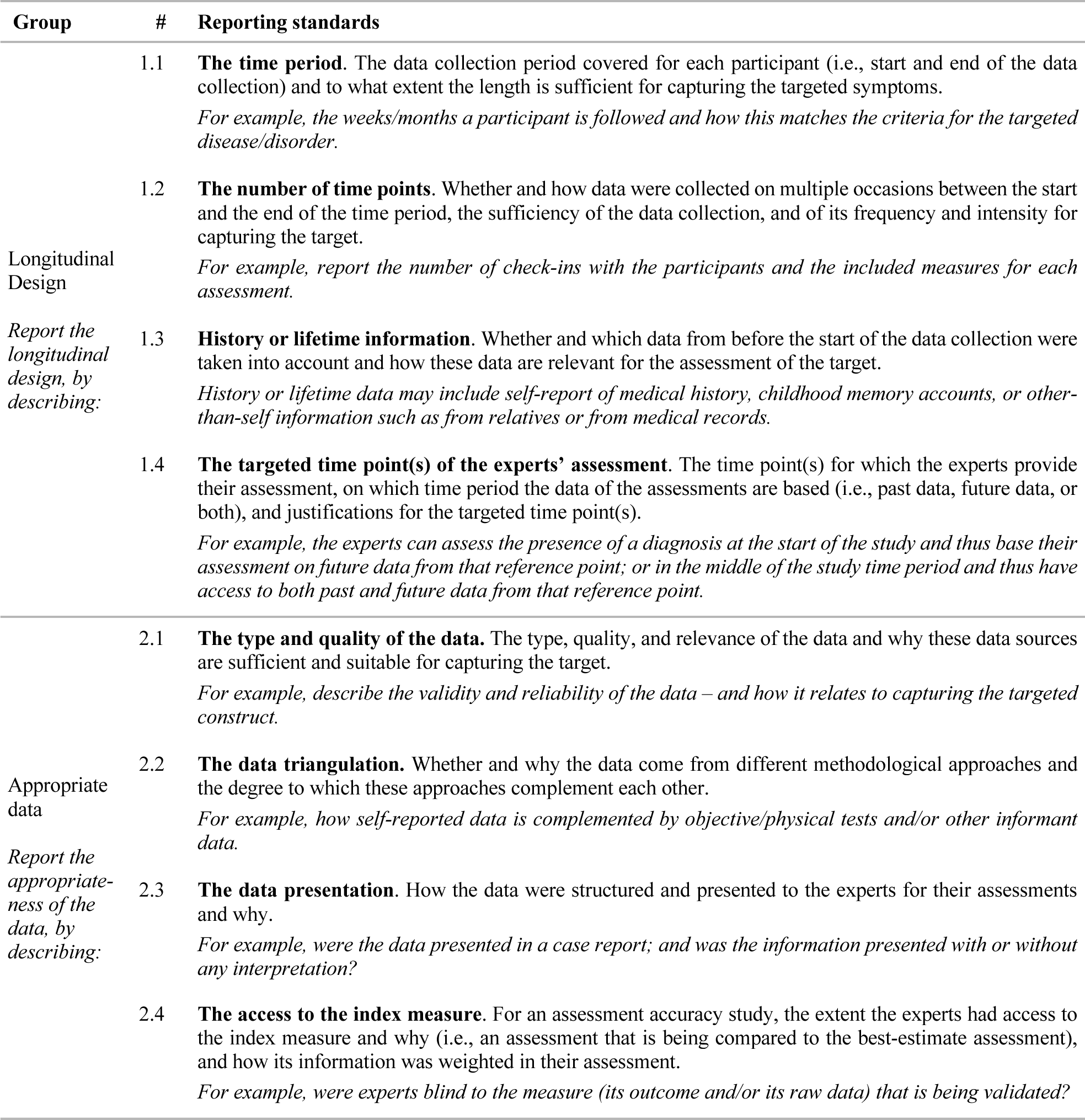
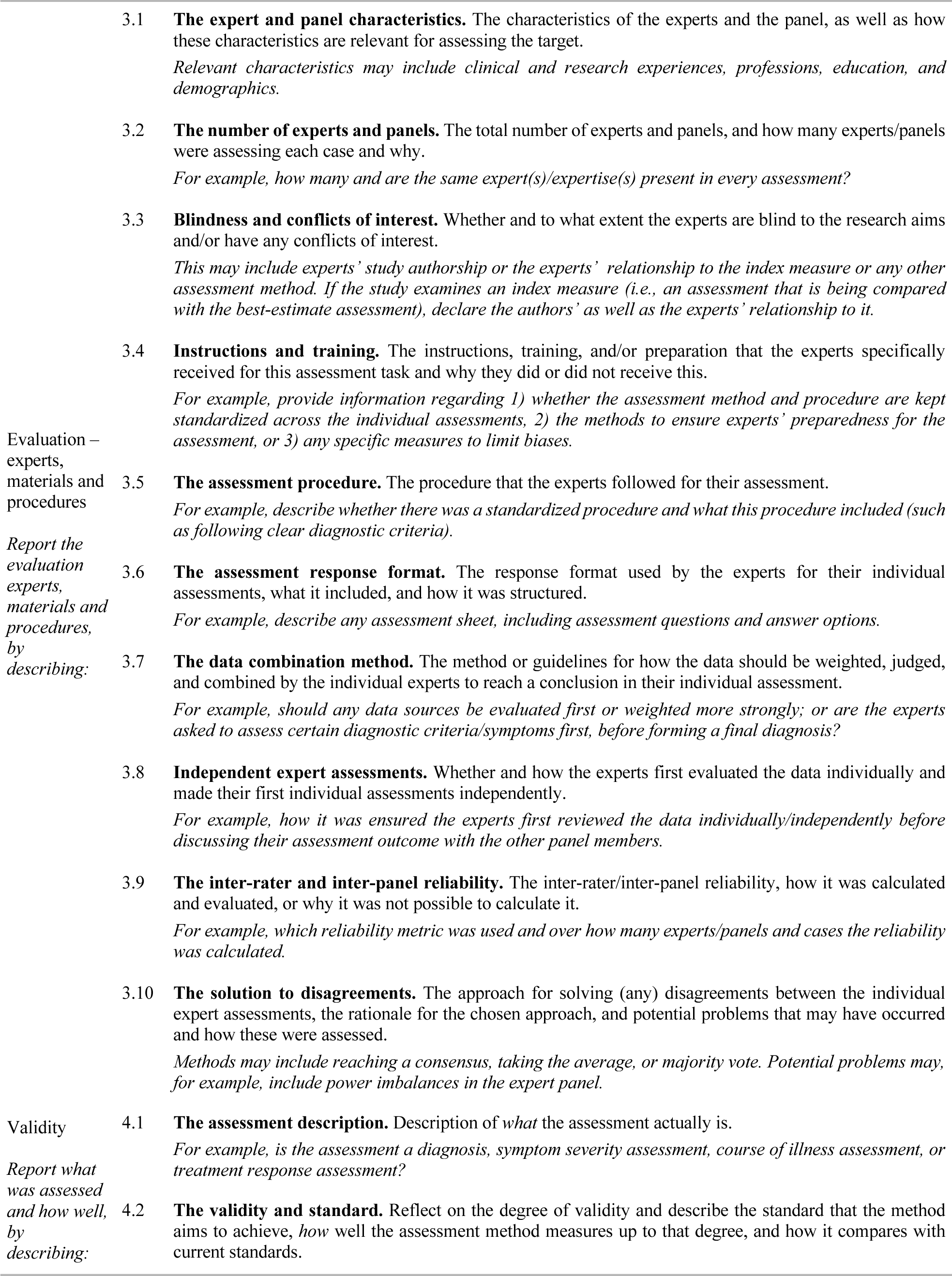
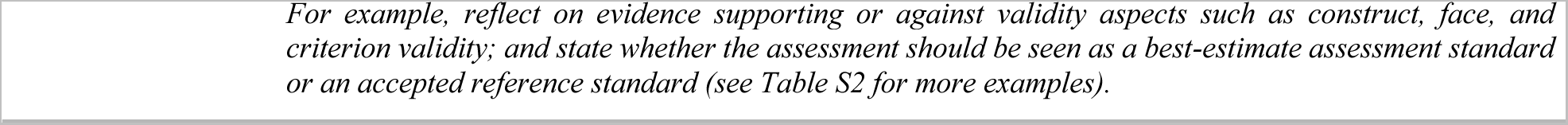
The LEADING guideline reporting standards.

| Group | # | Reporting standards |
| --- | --- | --- |
| Longitudinal Design<br>*Report the longitudinal design, by describing:* | 1.1 | **The time period.** The data collection period covered for each participant (i.e., start and end of the data collection) and to what extent the length is sufficient for capturing the targeted symptoms.<br>*For example, the weeks/months a participant is followed and how this matches the criteria for the targeted disease/disorder.* |
|  | 1.2 | **The number of time points.** Whether and how data were collected on multiple occasions between the start and the end of the time period, the sufficiency of the data collection, and of its frequency and intensity for capturing the target.<br>*For example, report the number of check-ins with the participants and the included measures for each assessment.* |
|  | 1.3 | **History or lifetime information.** Whether and which data from before the start of the data collection were taken into account and how these data are relevant for the assessment of the target.<br>*History or lifetime data may include self-report of medical history, childhood memory accounts, or other-than-self information such as from relatives or from medical records.* |
|  | 1.4 | **The targeted time point(s) of the experts' assessment.** The time point(s) for which the experts provide their assessment, on which time period the data of the assessments are based (i.e., past data, future data, or both), and justifications for the targeted time point(s).<br>*For example, the experts can assess the presence of a diagnosis at the start of the study and thus base their assessment on future data from that reference point; or in the middle of the study time period and thus have access to both past and future data from that reference point.* |
| Appropriate data<br>*Report the appropriateness of the data, by describing:* | 2.1 | **The type and quality of the data.** The type, quality, and relevance of the data and why these data sources are sufficient and suitable for capturing the target.<br>*For example, describe the validity and reliability of the data – and how it relates to capturing the targeted construct.* |
|  | 2.2 | **The data triangulation.** Whether and why the data come from different methodological approaches and the degree to which these approaches complement each other.<br>*For example, how self-reported data is complemented by objective/physical tests and/or other informant data.* |
|  | 2.3 | **The data presentation.** How the data were structured and presented to the experts for their assessments and why.<br>*For example, were the data presented in a case report; and was the information presented with or without any interpretation?* |
|  | 2.4 | **The access to the index measure.** For an assessment accuracy study, the extent the experts had access to the index measure and why (i.e., an assessment that is being compared to the best-estimate assessment), and how its information was weighted in their assessment.<br>*For example, were experts blind to the measure (its outcome and/or its raw data) that is being validated?* |
| Evaluation – experts, materials and procedures<br>Report the evaluation experts, materials and procedures, by describing: | 3.1 | **The expert and panel characteristics.** The characteristics of the experts and the panel, as well as how these characteristics are relevant for assessing the target.<br>*Relevant characteristics may include clinical and research experiences, professions, education, and demographics.* |
|  | 3.2 | **The number of experts and panels.** The total number of experts and panels, and how many experts/panels were assessing each case and why.<br>*For example, how many and are the same expert(s)/expertise(s) present in every assessment?* |
|  | 3.3 | **Blindness and conflicts of interest.** Whether and to what extent the experts are blind to the research aims and/or have any conflicts of interest.<br>*This may include experts' study authorship or the experts' relationship to the index measure or any other assessment method. If the study examines an index measure (i.e., an assessment that is being compared with the best-estimate assessment), declare the authors' as well as the experts' relationship to it.* |
|  | 3.4 | **Instructions and training.** The instructions, training, and/or preparation that the experts specifically received for this assessment task and why they did or did not receive this.<br>*For example, provide information regarding 1) whether the assessment method and procedure are kept standardized across the individual assessments, 2) the methods to ensure experts' preparedness for the assessment, or 3) any specific measures to limit biases.* |
|  | 3.5 | **The assessment procedure.** The procedure that the experts followed for their assessment.<br>*For example, describe whether there was a standardized procedure and what this procedure included (such as following clear diagnostic criteria).* |
|  | 3.6 | **The assessment response format.** The response format used by the experts for their individual assessments, what it included, and how it was structured.<br>*For example, describe any assessment sheet, including assessment questions and answer options.* |
|  | 3.7 | **The data combination method.** The method or guidelines for how the data should be weighted, judged, and combined by the individual experts to reach a conclusion in their individual assessment.<br>*For example, should any data sources be evaluated first or weighted more strongly; or are the experts asked to assess certain diagnostic criteria/symptoms first, before forming a final diagnosis?* |
|  | 3.8 | **Independent expert assessments.** Whether and how the experts first evaluated the data individually and made their first individual assessments independently.<br>*For example, how it was ensured the experts first reviewed the data individually/independently before discussing their assessment outcome with the other panel members.* |
|  | 3.9 | **The inter-rater and inter-panel reliability.** The inter-rater/inter-panel reliability, how it was calculated and evaluated, or why it was not possible to calculate it.<br>*For example, which reliability metric was used and over how many experts/panels and cases the reliability was calculated.* |
|  | 3.10 | **The solution to disagreements.** The approach for solving (any) disagreements between the individual expert assessments, the rationale for the chosen approach, and potential problems that may have occurred and how these were assessed.<br>*Methods may include reaching a consensus, taking the average, or majority vote. Potential problems may, for example, include power imbalances in the expert panel.* |
| Validity<br>Report what was assessed and how well, by describing: | 4.1 | **The assessment description.** Description of *what* the assessment actually is.<br>*For example, is the assessment a diagnosis, symptom severity assessment, course of illness assessment, or treatment response assessment?* |
|  | 4.2 | **The validity and standard.** Reflect on the degree of validity and describe the standard that the method aims to achieve, *how well* the assessment method measures up to that degree, and how it compares with current standards. |

*Instructions.* The LEADING guideline comprises these 20 reporting standards for comprehensive reporting of assessment methods involving expert(s) reviewing several sources of information (over time) to achieve a more accurate assessment (e.g., see Expert Panel, Best-Estimate diagnosis, and Longitudinal Expert All Data methods). The standards aim to help researchers plan, carry out, and report studies employing these assessment methods, as well as help readers evaluate them. As such, avoid simply answering yes or no to the standards when you instead can (succinctly) *describe* justifications and courses of action. Ensure the reports of the standards are clear, specific, and justified. To exemplify, standard *1.1 The time period* could be reported as ‘*The time span was six weeks, which covers more than the two weeks a person should have the symptoms for meeting the criteria for Major Depressive Disorder according to the DSM-5.*’

Not all of the reporting standards will be applicable to all types of studies – however, it is typically better to describe how a standard is not applicable than to leave the information out. Since the guideline covers the reporting of the assessment method, the method section would suit the reporting of most standards in most cases. However, the reporting guideline does *not* standardize where standards should be reported. When standards are considered less relevant or not applicable to a specific study, they can, for example, be described in an Appendix. Since the guideline focuses specifically on the reporting of the assessment method, it is recommended to use a complementary guideline for the reporting of the other study components: Which complementary guideline is dependent on the study type in which the assessment method is employed (e.g., see STARD for diagnostic accuracy studies; STROBE for observational studies; and CONSORT for randomized trials).

### Using the LEADING guideline to evaluate published studies

Evaluating a random selection of 30 articles indicates severe heterogeneity in *what* of the methods is reported and *how*, suggesting the need for a guideline that enables comprehensive reporting of these assessment methods (Table 2; see the SM for the search strategy). Across the 30 studies, 10 to 63% (Mean = 33%) of the standards were *not* reported. Regarding the reporting standards, the type and quality of the data (2.1), the access to the index measure (2.4), the expert and panel characteristics (3.1), the number of experts and panels (3.2), the assessment procedure (3.5), and the assessment description (4.1) were mostly reported (i.e., green in more than 50% of the studies). However, the data presentation (2.3), the instructions and training (3.4), the data combination method (3.7), the inter-rater and inter-panel reliability (3.9), and the validity and standard (4.2) were *not* reported at all in the majority of the studies (i.e., red in more than 50% of the studies). Considering that most changes that resulted from discussing disagreements between the raters (V.E. and K.K.) were from green to orange, and that green refers to a (minimally) sufficiently reported and orange to insufficiently reported, this suggests that the table is conservative in regards to the severity of the current state of poor reporting (i.e., potentially showing a more positive picture; for more information see the SM).

**Table 2.**
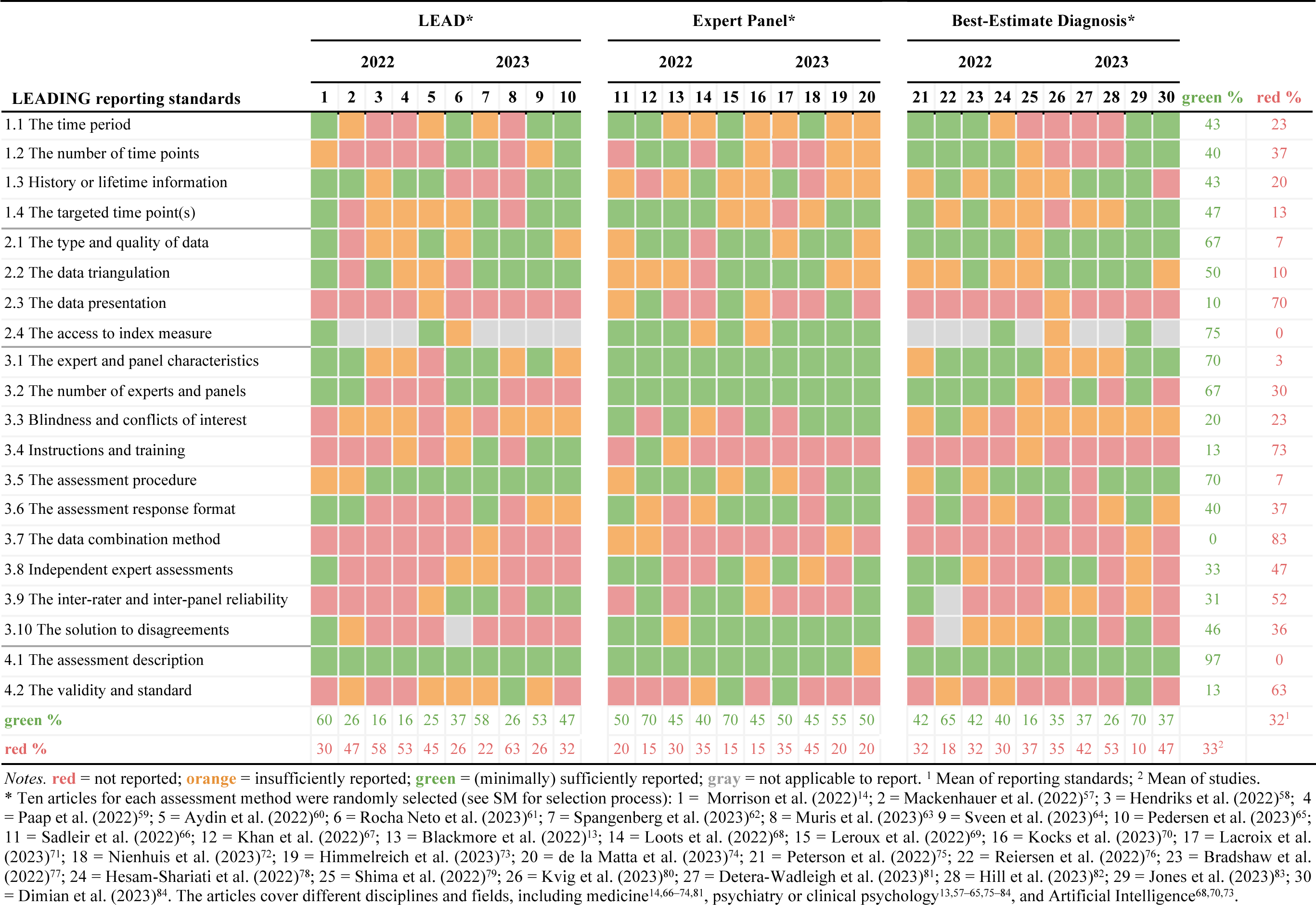
Reports across the LEADING guideline standards in 30 randomly selected articles published in 2022 and 2023.

## Discussion

Our objective was to develop a guideline that supports comprehensive reporting of studies collecting longitudinal, appropriate data that experts evaluate to achieve an assessment that is more accurate than a single error-prone measure. The aim is to help researchers plan, report, and evaluate the assessment method-related elements of this study design.

The LEADING reporting standards were established through an open process, incorporating relevant empirical evidence and methodological work, complementary reporting guidelines, and comprehensive iterations of expert feedback and patients’ perspectives. As this guideline focuses on the assessment methods, we recommend that researchers also rely on established guidelines for other parts of their research, such as sampling and other epidemiological aspects (e.g., STROBE^49^, CONSORT^51^, and STARD^50^). We encourage knowledge about and adherence to the LEADING guideline via scientific journals, editorials, and the EQUATOR network, as well as inclusion in research method courses in clinical studies.

### Limitations

We have connected three assessment methods with similar approaches from related fields and drafted applicable reporting standards. We presented the rationale for selecting these three methods and each reporting standard with supporting evidence in the Delphi survey for review, which did not bring up additional methods or reporting standards. However, as we did not carry out a systematic literature review of the three identified literature bodies or for each of the reporting standards, we cannot exclude the existence of other assessment methods with similar approaches: We welcome any suggestions about similar methods to which the guideline is applicable.

Although the Delphi survey participants and the author group had a wide range of experiences and backgrounds, psychiatry and clinical psychology (*n* = 18) were overrepresented as compared to, for example, other areas of medicine (*n* = 8) in the Delphi and author group. Geographically, Europe and North America were the most common in the Delphi and author group, whereas several areas were not represented. The Delphi participants were the first or last authors of studies employing the assessment methods. However, the quality of the articles, and the education or experience of the authors, were not taken into account as selection criteria (although it was self-reported as presented in Figure 1). Finally, the number of Delphi participants (27 in Round 1, 20 in Round 2) is relatively small compared to some other standard developments (e.g., 73 in the development of STARD^85^), but it is comparable to others (e.g., 24 for development of the TRIPOD statement^52^). Even though the response rate in Round 1 (21%) can be considered low, the number of participants was sufficient to cover a broad range of academic backgrounds, methodological experiences, and demographics (Figure 1). The same limitation is applicable to the size of the steering group (*n* = 7) as well as the test-user group (*n* = 2). The LEADING guideline should be regarded as an evolving reporting guideline requiring ongoing evaluation, refinement, and revision. Suggestions and recommendations for improvements are welcomed by emailing the corresponding authors.

### Conclusions

The LEADING guideline emphasizes the transparent reporting of the methodological components of the assessment method and the importance of reporting *what* was assessed and *how* well. Considering the increasing need for high-accuracy assessments in diverse fields, we hope that the LEADING guideline will be useful in assisting researchers in planning, carrying out, reporting, and evaluating research that aims to achieve accurate assessments.

## Supporting information

Supplementary_LEADING_Guideline

## Data Availability

Data sharing statement: Open data (Delphi surveys 1 and 2), code (analyses), and material (surveys) can be found on the Open Science Framework: https://osf.io/fkv4b/

## Declarations

### Ethics approval

Swedish law (2003:460) and the Swedish Ethical Review Authority state that only research that includes i) collecting personal information, and/or ii) that involves “obvious” (uppenbar) risk for physical or psychological harm, or iii) involves manipulating or deceiving individuals, should undergo an external ethical review. Considering that the current research involves asking participants to rate and comment on the reporting recommendations, this research does not fall within these criteria. The participants provided their informed consent and their individual open-ended comments and closed-ended ratings in the open material are anonymized. The closed-ended ratings per reporting standard in the Supplementary Material are presented on group level. The study is deemed exempt from requiring ethical approval according to Swedish Law (see §3-4 of the Act [2003:460] on ethical review of research involving humans in Sweden). Hence the research should not be reviewed by the Swedish Ethical Review Authority. More information can be found at the Swedish Ethical Review Authority (https://etikprovningsmyndigheten.se/).

### Availability of data and materials

Open data (Delphi rounds 1 and 2), code (analyses), and material (surveys) can be found on the Open Science Framework: https://osf.io/fkv4b/

## Authors’ contributions

The corresponding author attests that all listed authors meet authorship criteria and that no others meeting the criteria have been omitted.

*Contributor roles:* E = Equal; S = Supporting; L = Lead

*Working group:* V.C. Eijsbroek, K. Kjell, and O. Kjell.

*Steering group:* H.A. Schwartz, J.R. Boehnke, E.I. Fried, D.N. Klein, P. Gustafsson, I. Augenstein, and P.M.M. Bossuyt.

---

|  |  |
| --- | --- |
| Conceptualization | Working group |
| Data curation | V.E. |
| Formal analysis | V.E., O.K. (S) |
| Funding acquisition | O.K. (E) & K.K. (E) |
| Investigation | Working group |
| Methodology | Working group (L), Steering group |
| Project administration | Working group |
| Resources | Working group (L), Steering group |
| Software | - |
| Supervision | O.K. |
| Validation | - |
| Visualization | V.E. |
| Writing – original draft | Working group |
| Writing – review & editing | Working group (L), Steering group |
| Other | - |

## Acknowledgments

*Patient organization members*

E. Sellberg (Chairman of Libra Balans Skåne, board member of Balans National) and I. Odenbrand (Vice Chairman of Libra Balans Skåne)

*Participants in Delphi Round 1*

I. Augenstein (Professor, Computer Science, University of Copenhagen),

S. Aydin (PhD, Developmental and Educational Psychology, Leiden University),

E. Billstedt (Professor, Neuroscience and Physiology, University of Gothenburg),

J.R. Boehnke (PhD, School of Health Sciences, University of Dundee),

D.W. Black (MD, Professor, Medicine, University of Iowa Carver College of Medicine),

B. Cannell (Associate Professor, Public Health, University of Texas Health Science Center at Houston),

G.A. Carlson (Professor, Psychiatry, Stony Brook University),

K.A.S. Davis (Researcher, Psychiatry Psychology and Neuroscience, King’s College London),

F. Dereboy (MD, Psychiatry, Aydın Adnan Menderes University),

E.I. Fried (Associate Professor, Clinical Psychology, Leiden University),

P. Gustafsson (Associate Professor, Child and Adolescent Psychiatry, Lund University),

R. Handels (Assistant Professor, Psychiatry and Neuropsychology, Maastricht University),

K. Jenniskens (Assistant Professor, Clinical Epidemiology, Utrecht University),

C. Klaiman (Associate Professor, Pediatrics, Emory University),

D.N. Klein (Professor, Clinical Psychology, Stony Brook University),

M. McCloskey (Professor, Cognitive Science and Psychology, Johns Hopkins University),

A.C. Miers (Associate Professor, Developmental and Educational Psychology, Leiden University),

K.G.M. Moons (Professor, Clinical Epidemiology, Utrecht University),

L. Mosqueda (Professor, Medicine, University of Southern California),

H.A. Schwartz (Associate Professor, Computer Science, Stony Brook University),

M. Stein (Professor, Psychiatry and Behavioral Sciences, University of Washington),

J.G. Tillman (PhD, Clinical Psychology, Yale School of Medicine),

Y.P. Wang (MD, PhD, Medicine, University of Sao Paulo Medical School), and

J. Yonashiro-Cho (PhD, Medicine, University of Southern California).

*Participants in Delphi Round 2*

Augenstein (Professor, Computer Science, University of Copenhagen),

S. Aydin (PhD, Developmental and Educational Psychology, Leiden University),

J.R. Boehnke (PhD, School of Health Sciences, University of Dundee),

G.A. Carlson (Professor, Psychiatry, Stony Brook University),

K.A.S. Davis (Researcher, Psychiatry Psychology and Neuroscience, King’s College London),

E.I. Fried (Associate Professor, Clinical Psychology, Leiden University),

P. Gustafsson (Associate Professor, Child and adolescent psychiatry, Lund University),

R. Handels (Assistant Professor, Psychiatry and Neuropsychology, Maastricht University),

K. Jenniskens (Assistant Professor, Clinical Epidemiology, Utrecht University),

C. Klaiman (Associate Professor, Pediatrics, Emory University),

D.N. Klein (Professor, Clinical Psychology, Stony Brook University),

M. McCloskey (Professor, Cognitive Science and Psychology, Johns Hopkins University),

A.C. Miers (Associate Professor, Developmental and Educational Psychology, Leiden University),

K.G.M. Moons (Professor, Clinical Epidemiology, Utrecht University),

L. Mosqueda (Professor, Medicine, University of Southern California),

Y.P. Wang (MD, PhD, Medicine, University of Sao Paulo Medical School), and

J. Yonashiro-Cho (PhD, Medicine, University of Southern California).

*Test-users*

T. Ivarsson (Associate Professor, Neuroscience and Physiology, University of Gothenburg) and

P.J. Snelling (PhD, Medicine, Gold Coast University Hospital and Griffith University)

## Patient and public involvement

Prior to the Delphi procedure and test procedures, the reporting standards with inclusion rationales were discussed in an online workshop with a patient organization for depression (the chairman and vice chairman from Libra Balans Skåne), followed by receiving feedback from the steering group members to receive a wide range of perspectives early in the process.

