## Supplementary_LEADING_Guideline for "The LEADING Guideline. Reporting Standards for Expert Panel, Best-Estimate Diagnosis, and Longitudinal Expert All Data (LEAD) Studies"

### Supplementary Material

**Reporting guidelines and systematic reviews**

Beyond the review of expert panels applications[1]; the *STrengthening the Reporting of OBservational studies in Epidemiology statement* (STROBE)[2]; the *Statement for Reporting Diagnostic Accuracy Studies* (STARD)[3]; and the guidance for developers of health research reporting guidelines[4], other reporting guidelines and systematic reviews that were considered in the development of the LEADING reporting standards include:

- Optimizing the use of expert panel reference diagnoses in diagnostic studies of multidimensional syndromes[5];
- The *Transparent reporting of a multivariable prediction model for individual prognosis or diagnosis* (TRIPOD) statement[6];
- The *Consolidated Standards of Reporting Trials* (CONSORT)[7];
- The *Template for Intervention Description and Replication* (TIDieR) guidelines[8];
- Guidance on *Conducting and REporting DElphi Studies* (CREDES) in palliative care[9];
- The standards for reporting of core outcome sets (COS-STAR) statement[10];
- The reporting guideline for the early-stage clinical evaluation of decision support systems driven by artificial intelligence (DECIDE-AI)[11];
- A review of solutions for diagnostic accuracy studies with an imperfect or missing reference standard[12];
- the *Consolidated Health Economic Evaluation Reporting Standards* (CHEERS) statement[13]
- A methodological review for reporting of Delphi studies in health sciences[14]; and
- The *Checklist for the reporting of updated guidelines* (CheckUp)[15].

References, including all authors, are included in the reference list.

**Guideline development steps**

Table S1 presents the recommended steps for developing a health research reporting guideline[4] and a description of whether and how we followed each step and why.

**Table S1 | *Elaboration on the steps for developing a health research reporting guideline***

| Step | # | Description |
|---|---|---|
| *Initial steps* | 1 | **Identify the need for a guideline.**<br><br>The inconsistent and incomplete reporting of the assessment methods in Expert Panel, Best-Estimate Diagnosis, and LEAD studies shows the need for this guideline. These assessment methods claim to achieve best-estimate reference standards. However, the quality of such proclaimed best-estimate assessments varies substantially and is typically very difficult to evaluate due to poor reporting of the method on how they were achieved (e.g., see Table 2). |
| | 1.1 | **Develop new guidance.** |

Although a systematic review of assessment methods and reporting of expert panels identified and structured the various choices involved in Expert Panel studies, no attempt was made to develop a formal guideline for reporting such studies. The STARD statement (*Standards for Reporting of Diagnostic Accuracy Studies*) was developed to improve the completeness and transparency of reports of diagnostic accuracy studies; however, the applications of (best-estimate) assessments range much further than this design.

1.2 **Extend existing guidance.**

The reporting standards of previous guidelines are not sufficient for the complete reporting of the assessment methods where multiple experts review several sources of (longitudinal) information to form a best-estimate assessment. By specifically focusing on the reporting on the assessment method, the LEADING guideline aims to complement rather than repeat previous guidelines. The LEADING guideline does not standardize which other guideline should be used for reporting other study components since this depends on the type of study (e.g., see STARD for diagnostic accuracy studies, CONSORT for randomized controlled trials, and STROBE for observational studies).

1.3 **Implement existing guidance.**

This guideline does not include the implementation of an existing guideline in another field or clinical specialty (see 1.1 and 1.2)

---

2 **Review the literature.** See 2.1 to 2.3.

2.1 **Identify previous relevant guidance.**

In the first of the four development stages, relevant reporting guidelines and systematic reviews were identified, including a review of expert panel applications, the STROBE statement, and the STARD statement. Other complementary reporting guidelines and systematic reviews are presented in the supplementary material.

2.2 **Seek relevant guidance on the quality of reporting in published research articles.**

We connect three bodies of literature that employ silmiar assessment methods (Expert Panel, Best-Estimate Diagnosis, and LEAD) and the working group identified relevant research using or describing the assessment methods. For example, a systematic review of assessment methods and reporting of expert panels has demonstrated that the methods used for panel or consensus diagnoses vary substantially across studies and that many aspects of the procedure are often unclear or not reported at all. To further examine the reporting quality, V.E. and K.K. reviewed the reporting of the assessment methods in articles published in 2022 and 2023, which indicated severe heterogeneity in what parts of the methods and how these are reported.

2.3 **Identify key information related to the potential sources of bias in such studies.**

The assessment methods' procedure can be divided into four groups regarding the *longitudinal design*, the *data*, the *experts' evaluation*, and the *validity* of the assessment. We identified specific sources of bias for each group (see the Explanation and Elaboration including Table S4). The experts' evaluation especially includes many potential sources of biases since human decision-making is prone to cognitive biases as well as limited cognitive resources for the integration of a high amount of data sources. This group therefore includes the most reporting standards.

---

3 **Obtain funding for the guideline initiative.**

The project was funded by the funding of V.C. Eijsbroek, K. Kjell, and O. Kjell who received from FORTE (2022-01022).

---

Pre-meeting

4 **Identify participants.**

A working group (V.E., K.K., & O.K.) and a steering group (H.A.S., J.B., E.F., D.K., P.G., I.A., & P.B.) were formed to provide a wide range of expertise. The steering group included seven experts

| | | |
|---|---|---|
| | | and was selected to cover a diverse range of expertise and fields related to the assessment methods (e.g., psychiatry/clinical psychology, medicine, epidemiology, and Artificial Intelligence). The working group drafted the reporting standards. The standards were further developed with a patient organization for depression, followed by feedback from the steering group. |
| | 5 | **Conduct a Delphi exercise.**<br><br>To systematically collect expert feedback from different perspectives, we conducted a Delphi exercise. We used an iterative process based on two rounds of Delphi surveys, enabling feedback from round 1 to feed into round 2. The first and/or last authors of articles since 2013 using any of the three assessment methods as well as the seven steering group members, were invited to participate in Round 1. Only participants from Round 1 were invited to Round 2. No third Delphi round was carried out since i) the median agreement for each reporting standard in both rounds ranged from *Agree* to *Strongly Agree*, ii) no new standards were suggested, and iii) only minor changes were needed after Round 2. |
| | 6 | **Generate a list of items for consideration at the face-to-face meeting.**<br><br>Potential standards were drafted by the working group with the objective of comprehensive reporting of the assessment methods. The standards with inclusion rationales were further developed through a workshop with a patient organization for depression, followed by feedback from the steering group members. Subsequently, delphi participants provided feedback on the standards in the two Delphi surveys through open- and closed-ended response formats. The resulting standards were considered and discussed in an online meeting with the working group and steering group. |
| | 7 | **Prepare the face-to-face meeting.** See 7.1 to 7.5. |
| | 7.1 | **Decide the size and duration of the face-to-face meeting.**<br><br>All working group and steering group members were invited to the expert consensus meeting. Based on availabilities, the meeting date was scheduled (April 12th, 2023, 3-4 pm). All members except for one (K.K.) could attend ($N = 9$). |
| | 7.2 | **Develop meeting logistics.**<br><br>The expert consensus meeting was decided to be held online to make it possible for members from different continents to attend. |
| | 7.3 | **Develop meeting agenda.**<br><br>The content and structure of the consensus meeting were prepared by the working group (V.E., K.K., O.K.). |
| | 7.3.1 | **Consider presentations on relevant background topics, including a summary of evidence.**<br><br>The consensus meeting included going through the draft of the paper, including background information, rationales (explanation and elaboration), and aims of the guidelines, as well as of the individual reporting standards and groups. |
| | 7.3.2 | **Plan to share results of the Delphi exercise, if done.**<br><br>The consensus meeting included going through the findings of Delphi Rounds 1 and 2 and discussing their implications for the guideline and the individual reporting standards. Delphi results could be accessed in the main draft and supplementary of the paper in combination with raw materials and results on the Open Science Framework (OSF). |
| | 7.3.3 | **Invite session chairs.**<br><br>The meeting was led by the last author (O.K.) and was not divided into separate sessions. |

| | | |
|---|---|---|
| | 7.4 | **Prepare materials to be sent to participants prior to the meeting.**<br><br>Participants had access to the guideline including the individual reporting standards and groups with their elaboration and explanation (inclusion) rationales. The guideline and a drafted paper, including background information, rationals, and aims were sent to the participants two weeks before the consensus meeting. The participants had the option to provide comments and feedback in writing prior to the meeting. |
| | 7.5 | **Arrange to record the meeting.**<br><br>The meeting was not recorded. |
| Meeting | 8 | **Present and discuss the results of pre-meeting activities and relevant evidence.**<br><br>The draft was presented and discussed, including background information, rationales, and aims of the guidelines. Also, the individual reporting standards and groups were discussed, including their background information and inclusion rationales (explanations and elaborations). Finally, the Delphi results and their implications were discussed. All participants agreed on not conducting a third Delphi round. |
| | 8.1 | **Discuss the rationale for including items in the checklist.**<br><br>The criteria for including a reporting standard was that the median of Delphi expert responses was at least *Agree* on the question about its inclusion. A couple of reporting standard formulations were clarified based on discussions in the consensus meeting, such as what we mean by *validity* and *standard* in the last reporting standard group. |
| | 8.2 | **Discuss the development of a flow diagram.**<br><br>The development of a flow diagram was not discussed, since the number of standards included in the guideline did not change a lot during the Delphi procedure or consensus discussion. One standard was removed after Delphi Round 1; otherwise the changes consisted of clarifications and reformulations. |
| | 8.3 | **Discuss strategy for producing documents; identify who will be involved in which activities; discuss authorship.**<br><br>During the consensus meeting, it was decided to produce one instead of several documents. Explanations and elaborations (e.g., inclusions, rationales, and examples) will be presented in the main draft in combination with the supplementary. |
| | 8.4 | **Discuss knowledge translation strategy.**<br><br>Potential venues for publication were discussed including different journals as well as editorials, and the EQUATOR network. |
| Post-meeting | 9 | **Develop the guidance statement.**<br><br>Based on the consensus meeting and the Delphi findings, the guideline was further developed by the working group in iterations with the steering group. The guideline was developed and structured, including an introduction (i.e., information about the assessment methods and reporting standards), method section (i.e., the development of the guideline), result section (i.e., the guideline's reporting standards and groups), and discussion (i.e., implications and limitations of the guideline). |
| | 9.1 | **Pilot test the checklist.**<br><br>The guideline was piloted/tested i) by researchers with experience of each assessment method answering and/or commenting on each reporting standard, and ii) by the authors (V.E., K.K.) using it to evaluate published articles. First, two independent researchers reported each standard and/or provided feedback about the formulation of the standards in a survey. Second, V.E. and K.K. |

| | | |
|---|---|---|
| | | evaluated the reporting of the standards in fifteen articles published in 2022 and 2023. Both test approaches resulted in clarifications in the descriptions and examples of the reporting standards. |
| | 10 | **Develop an explanatory document (E&E).**<br><br>No separate explanatory document was developed. The individual reporting standards, including examples for each standard, are presented in the table in the main draft. Inclusion rationales and evidential support for each of the individual standards and the four groups are presented in the supplementary material. |
| | 11 | **Develop a publication strategy.**<br><br>A list of potential venues was made for submission and publication of the guideline. After publication, the strategy includes promotion in editorials as well as publishing it on the EQUATOR network. Also, potential inclusion into research method courses in clinical studies, such as medicine and psychology, is considered. |
| | 11.1 | **Consider multiple and simultaneous publications.**<br><br>The guideline is currently in the stage of first submission and publication. Publication in other journals and on the EQUATOR network as well as recommendations in editorials, will be discussed after. |
| Post-publication | 12 | **Seek and deal with feedback and criticism.** |
| | 13 | **Encourage guideline endorsement.** |
| | 14 | **Support adherence to the guideline.** |
| | 15 | **Evaluate the impact of the reporting guidance.** |
| | 16 | **Develop Web site.** |
| | 17 | **Translate guideline.** |
| | 18 | **Update guideline.** |

### Search strategies

Articles using any of the three assessment methods were identified through a literature search using Google Scholar with the following search terms: [“expert panel diagnosis” OR “expert panel assessment” OR “expert panel consensus” OR “expert panel methodology” OR “expert panel standard” OR “expert panel reference”] for Expert Panel studies; [“best-estimate diagnosis” OR “best-estimate diagnostic” OR “best-estimate standard” OR “best-estimate assessment” OR “best-estimate methodology” OR “best-estimate reference”] for Best-Estimate Diagnosis studies; and [“longitudinal expert all data” OR “longitudinal evaluation all data”] for LEAD studies. Articles that clearly stated the use of one of the three assessment methods were selected. Articles stating another purpose than assessment were excluded (e.g. when an expert panel was used to reach a consensus about a treatment strategy).

*Delphi participants*. The first and last authors of articles since 2013 that include a LEAD, Expert Panel, or Best-Estimate Diagnosis method were invited to the Delphi surveys (as well as the steering group members). Articles were excluded when the email address of the first and last authors could not be obtained. The result was an extensive (but not exhaustive) list of 87 articles, including 41 LEAD studies, 21 Expert Panel studies, and 25 Best-Estimate Diagnosis studies from which the first and/or last author was emailed (See the Delphi reference list[1-87]). Some authors forwarded the survey to co-authors or colleagues who made use of one of the assessment methods in their research.

*Test-users*. Test-user feedback was collected (i.e., piloting the reporting guideline) via an online test-user survey. The test users ($n$ = 2) were the first or last author of a LEAD, Expert Panel, or Best Estimate Diagnosis study published in 2023 or recruited via networks of the authors. They received an introduction to the reporting guideline and were instructed to report each of the 20 standards based on a planned, ongoing, or finished study they are involved in using a LEAD, Best-Estimate Diagnosis, or Expert Panel method. Additionally, they had the possibility to give feedback on the formulations of the reporting standards via open-ended questions.

*Reports of the standards in 2022*. Two authors (V.E., K.K.) examined which standards are reported and how well in 30 articles published in 2022 and 2023. Articles that referred to a previous article for reporting of the assessment method were excluded. From each year, the first 20 articles were selected in the Google Scholar searches for Expert Panel and Best-Estimate Diagnosis studies, of which five from each method from each year were randomly selected for reviewing the reports of the standards. The Google Scholar search for LEAD studies generated ten articles published in 2022 and 11 articles published in 2023, of which five from each year were randomly selected for reviewing the reports. See the Standard reporting references list for 2022[1-50] and 2023[1-51] of which the reviewed articles are underlined.

#### Delphi survey results

Tables S1 and S2 present the results from the closed-ended ratings for each reporting standard from the Delphi surveys. The Delphi surveys, as well as the anonymized open-ended results and individual closed-ended ratings, can be found in the open material (https://osf.io/fkv4b/).

**Table S2 | *Reporting standards and ratings Delphi Round 1***

| | | | *This item should be included in the reporting checklist (1 - 7)* | | | *Whether this information is present or not would influence my perceptions of the quality of a study (1 - 7)* | | |
|---|---|---|---|---|---|---|---|---|
| **Group** | **#** | **Reporting standards** | **Mean (SD)** | **Median** | **Min - Max (Range)** | **Mean (SD)** | **Median** | **Min - Max (Range)** |
| Longitudinal Design | 1.1 | Time period | 6.3 (0.6) | 6 | 5 - 7 (2) | 5.6 (1.2) | 6 | 3 - 7 (4) |
| | 1.2 | Number of time points | 6.2 (0.9) | 6 | 4 - 7 (3) | 5.7 (1.4) | 6 | 2 - 7 (5) |
| | 1.3 | Retrospective account | 5.6 (1.6) | 6 | 1 - 7 (6) | 5.1 (1.5) | 6 | 1 - 7 (6) |
| | 1.4 | Targeted time point(s) of the expert assessment | 6.2 (0.9) | 6 | 4 - 7 (3) | 5.7 (1.2) | 6 | 2 - 7 (5) |
| Evaluation - Expert and Procedure | 2.1 | Expert characteristics and experiences | 6.3 (1.0) | 7 | 4 - 7 (3) | 5.6 (1.6) | 6 | 1 - 7 (6) |
| | 2.2 | Blindness and Conflicts of Interest | 6.3 (1.1) | 7 | 3 - 7 (4) | 5.6 (1.5) | 6 | 1 - 7 (6) |

| | | | | | | | | |
|---|---|---|---|---|---|---|---|---|
| | 2.3 | Instructions and training | 5.9 (1.5) | 7 | 2 - 7 (5) | 5.4 (1.6) | 6 | 1 - 7 (6) |
| | 2.4 | Assessment criteria | 6.3 (0.9) | 7 | 4 - 7 (3) | 5.9 (1.4) | 6 | 1 - 7 (6) |
| | 2.5 | Assessment sheet | 5.6 (1.2) | 6 | 2 - 7 (5) | 5.2 (1.7) | 5 | 1 - 7 (6) |
| | 2.6 | Data combination method | 6.1 (0.9) | 6 | 4 - 7 (3) | 5.5 (1.3) | 6 | 2 - 7 (5) |
| | 2.7 | Number of experts | 6.2 (1.1) | 6 | 3 - 7 (4) | 5.8 (1.3) | 6 | 2 - 7 (5) |
| | 2.8 | Independent assessments | 6.3 (1.0) | 7 | 4 - 7 (3) | 5.9 (1.4) | 6 | 1 - 7 (6) |
| | 2.9 | Inter-rater reliability | 5.9 (1.2) | 6 | 3 - 7 (4) | 5.7 (1.3) | 6 | 2 - 7 (5) |
| | 2.10 | Solution to disagreements | 6.3 (0.9) | 7 | 4 - 7 (3) | 6.0 (1.2) | 6 | 2 - 7 (5) |
| Apt Data | 3.1 | Type and quality of data | 6.2 (1.0) | 6 | 3 - 7 (4) | 5.9 (1.4) | 6 | 1 - 7 (6) |
| | 3.2 | Triangulation and multimodal data fusion | 6.1 (1.0) | 6 | 4 - 7 (3) | 5.6 (1.5) | 6 | 1 - 7 (6) |
| | 3.3 | Presentation: case report | 5.8 (1.2) | 6 | 3 - 7 (4) | 5.2 (1.3) | 5 | 2 - 7 (5) |
| | 3.4 | Access to the index measure | 6.0 (1.1) | 6 | 4 - 7 (3) | 5.7 (1.3) | 6 | 2 - 7 (5) |
| Assessment Validity | 4.1 | Description of the assessment | 6.7 (0.7) | 7 | 4 - 7 (3) | 6.2 (1.3) | 7 | 1 - 7 (6) |
| | 4.2 | Type/degree of standard | 5.9 (1.1) | 6 | 4 - 7 (3) | 5.4 (1.3) | 6 | 2 - 7 (5) |
| | 4.3 | Transparency and replicability | 5.4 (1.4) | 6 | 2 - 7 (5) | 5.3 (1.3) | 5 | 3 - 7 (4) |

### Table S3 | *Reporting standards and ratings Delphi Round 2*

| | | | *This item should be included in the reporting checklist (1 - 7)* | | | *Whether this information is present or not would influence my perceptions of the quality of a study (1 - 7)* | | |
|---|---|---|---|---|---|---|---|---|
| **Group** | **#** | **Reporting standards** | **Mean (SD)** | **Median** | **Min - Max (Range)** | **Mean (SD)** | **Median** | **Min - Max (Range)** |
| Longitudinal Design | 1.1 | The time period | 6.5 (0.6) | 7 | 5 - 7 (2) | 5.8 (1.3) | 6 | 2 - 7 (5) |
| | 1.2 | The number of time points | 6.4 (0.6) | 6 | 5 - 7 (2) | 5.7 (1.3) | 6 | 2 - 7 (5) |
| | 1.3 | Retrospective accounts | 5.8 (0.8) | 6 | 4 - 7 (3) | 5.3 (1.2) | 5 | 3 - 7 (4) |
| | 1.4 | The targeted time point(s) of the experts' assessment | 6.1 (1.2) | 6 | 2 - 7 (5) | 5.3 (1.4) | 6 | 2 - 7 (5) |
| Evaluation - Expert and Procedure | 2.1 | The expert and panel characteristics | 6.5 (0.6) | 7 | 5 - 7 (2) | 5.6 (1.5) | 6 | 1 - 7 (6) |
| | 2.2 | The number of experts and panels | 6.4 (0.8) | 6.5 | 5 - 7 (2) | 5.5 (1.2) | 5.5 | 2 - 7 (5) |
| | 2.3 | Blindness and conflicts of interests | 6.5 (1.2) | 7 | 2 - 7 (5) | 5.6 (1.9) | 6 | 1 - 7 (6) |
| | 2.4 | Instructions and training | 6.5 (0.5) | 6 | 6 - 7 (1) | 5.6 (1.2) | 6 | 2 - 7 (5) |
| | 2.5 | The assessment procedure | 6.6 (0.6) | 7 | 5 - 7 (2) | 5.8 (1.6) | 6 | 1 - 7 (6) |
| | 2.6 | The assessment response format | 5.8 (0.9) | 6 | 4 - 7 (3) | 4.9 (1.2) | 5 | 2 - 7 (5) |
| | 2.7 | The data combination method | 6.2 (1.0) | 6 | 3 - 7 (4) | 5.3 (1.7) | 6 | 1 - 7 (6) |

| | | | | | | | | |
|---|---|---|---|---|---|---|---|---|
| | 2.8 | Independent expert assessments | 6.5 (0.6) | 6.5 | 5 - 7 (2) | 5.5 (1.6) | 6 | 1 - 7 (6) |
| | 2.9 | The inter-rater and inter-panel reliability | 6.2 (0.5) | 6 | 5 - 7 (2) | 5.4 (1.6) | 6 | 2 - 7 (5) |
| | 2.10 | The solution to disagreements | 6.5 (0.5) | 6.5 | 6 - 7 (1) | 5.7 (1.6) | 6 | 1 - 7 (6) |
| Appropriate Data | 3.1 | The type and quality of the data | 6.3 (0.7) | 6 | 4 - 7 (3) | 5.8 (1.5) | 6 | 2 - 7 (5) |
| | 3.2 | Triangulation and multimodal data fusion | 5.5 (1.1) | 6 | 2 - 7 (5) | 5.0 (1.3) | 5 | 2 - 6 (4) |
| | 3.3 | The data presentation | 5.6 (0.9) | 6 | 4 - 7 (3) | 4.7 (1.2) | 5 | 2 - 7 (5) |
| | 3.4 | The access to the index measure | 6.1 (0.9) | 6 | 4 - 7 (3) | 5.3 (1.6) | 5.5 | 1 - 7 (6) |
| Assessment Validity | 4.1 | The assessment | 6.7 (0.7) | 7 | 4 - 7 (3) | 6.1 (1.9) | 7 | 1 - 7 (6) |
| | 4.2 | The degree of standard | 5.5 (1.4) | 6 | 2 - 7 (5) | 4.9 (1.6) | 5 | 2 - 7 (5) |

**Explanation and Elaboration**

Below, we describe the content of the LEADING guideline in more detail. The LEADING reporting standards are divided into four groups: *1. The longitudinal design group*, *2. The appropriate data group*, *3. The evaluation – experts, materials, and procedures group*, and *4. The validity group*. Elaborations on inclusion rationales and evidential support for each group are presented below, and evidential support for each individual reporting standard is presented in Table S4.

***1. The longitudinal design group*** consists of four reporting standards describing to what extent the assessment method includes longitudinal data (e.g., the data collection period and the number of time points), or is limited to a single examination performed at one point in time.

Many diagnostic criteria are longitudinal and differ across disorders (e.g., criteria specify a two-week period for Major Depressive Disorder versus a two-year period for Persistent Depressive Disorder[16]). Hence, the time period that the data cover should be sufficiently longitudinal and fine-grained to capture the targeted symptoms so that "symptoms that only emerge or are identified after an initial evaluation are also taken into account"[17] (*p*. 409). In other words, the time period represented by the data should reflect the specific symptoms targeted in the study.

It is difficult to perfectly recall symptoms retrospectively, and recall can be biased, for example, by the patient's current mood[18,19]. Prospective longitudinal studies[20], including Ecological Momentary Assessment methods[21], allow patients to repeatedly report their thoughts, feelings, and behaviors in real-time and in their natural settings, resulting in a more valid and dynamic representation over time and across contexts[22,23].

***2. The appropriate data group*** comprises four standards reporting to what extent the data are suitable for the assessment task (e.g., data quality, data sources/triangulation, and presentation of the data).

It is important to describe which data are used (e.g., quality, diversity, relevance, and sufficiency) and how it is being presented (e.g., structure and format). The LEADING guideline emphasizes a focus on the *quality* instead of just the *quantity* of the data (c.f., *All Data* in LEAD[17]).

Using data from diverse sources and different methodological approaches with differing sources of biases is called triangulation or multimodal methods/data fusion; these methods can be used to improve causal inference[24–27]. Since every single measure in psychology can be seen as fallible in some respect[28,29], triangulation rather than using a single type of measurement is important. However, considering human limitations in cognitive resources and cognitive biases, too much data can result in overload. When the amount of information becomes too much or too complex, it is harder to weigh and judge all data and make an accurate decision[30,31]. Therefore, it is important to consider the *appropriateness* of the data (rather than just the quantity) and how it is presented.

***3. The evaluation – experts, materials, and procedures group*** comprises 10 reporting standards addressing the need to clearly report the evaluation of the data, including both the *experts* carrying out the assessments (e.g., number of experts, characteristics, blindness) as well as any assessment *materials* and *procedures* (e.g., instructions, assessment criteria, response format, and agreement procedure).

Humans' decisions are fallible, including limited cognitive resources and biases – in this respect, the evaluation includes subjective interpretations. Therefore, it is important to describe the evaluation in great detail, such as how the data are judged, weighted, and combined in a systematic way[32] and how disagreements between the experts are resolved (e.g., how consensus is reached and how power imbalances in panels are handled[33]).

There are many different cognitive biases, including confirmatory strategies, premature closure, and insufficient use of base rates[34,35]. These biases can influence clinical decision-making and adversely impact the accuracy of clinical judgments[34–36]. In addition, the human brain has limited cognitive resources to take into account large amounts of data for inferential purposes[30]. Bowes et al.[34] state that "decreasing reliance on working memory and bolstering psychoeducation [on, e.g., cognitive biases] may alleviate the effects of bias on clinical decision-making" (*p.* 435).

***4. The validity group*** comprises two reporting standards addressing the need to clearly describe *what* was measured and *how* well.

A best-estimate assessment should live up to being the best available measure at a certain point in time[37,38]. Best-estimate assessments can constantly be challenged and superseded[29]. To develop and improve best-estimate assessments and to facilitate the processes of challenging them, the validity of the assessment and the assessment standards should clearly be described (see examples in Table S5). Logically, validation starts with a statement of the interpretation of an assessment, which includes specifying what the assessment intends to measure[39]. Validation further involves evaluating arguments in support of and against the intended interpretation of the assessment. There are several different validity aspects to consider (e.g., construct and criterion validity) that are of key relevance for the assessment and its purpose.

In addition, the very aim of these assessment methods is to improve accuracy in the absence of a single error-free measure – it is, thus, central to clearly define and reflect on the achieved validity and standard. This clarity addresses the problem that researchers sometimes refer to assessments using these methods as being *the gold standard* without describing what they mean by that term and how the assessment method may have succeeded in achieving that. Clearly describing *what* was measured and *how* well encourages authors to transparently and critically evaluate their assessment method and to compare (contextualize) it with other research and methods, which in turn further helps readers' critical evaluation.

***Verisimilitude: Degree of standard.*** The *best-estimate assessment* is not the absolute truth – it is highly dependent on the assessment method, and because it can be challenged and superseded, we consider it best conceptualized as a matter of *degree*. Popper's[40] term *verisimilitude*, or *truthlikeness*, is the notion that some propositions or theories are closer to the absolute truth than others whilst still being considered false; this corresponds to the best-estimate assessment standard. Similarly, assessment results can be compared to the extent they have been derived from the assessment method that is most likely to yield the most *truthlike* results.

**Table S4 |** ***The LEADING guideline reporting standards with evidential support***

| Group | # | Reporting standards |
|---|---|---|
| Longitudinal Design<br>*Report the longitudinal design, by describing:* | 1.1 | **The time period**. The data collection period covered for each participant (i.e., start and end of the data collection) and to what extent the length is sufficient for capturing the targeted symptoms.<br>*Main reasons:* Diagnostic and symptoms criteria are often longitudinal, and the period often differs across targets (e.g., see [16]). |
| | 1.2 | **The number of time points**. Whether and how data were collected on multiple occasions between the start and the end of the time period, the sufficiency of the data collection, and its frequency and intensity for capturing the target.<br>*Main reasons:* Symptoms are dynamic and most accurately assessed in real-time using, for example, ecological momentary assessment methods[20,22,23]. |
| | 1.3 | **History or lifetime information**. Whether and which data from before the start of the data collection were taken into account and how these data are relevant for the assessment of the target.<br>*Main reasons:* History or lifetime data provide context and can place the other data in perspective[41–43], but it can also be subject to, for example, recall biases[18,19]. |
| | 1.4 | **The targeted time point(s) of the experts' assessment**. The time point(s) for which the experts provide their assessment, on which time period the data of the assessments are based (i.e., past data, future data, or both), and justifications for the targeted time point(s).<br>*Main reasons:* Symptoms change over time[44,45], so it should be clear which point in time the experts' assessments refer to. |
| Appropriate data<br>*Report the appropriate-ness of the data, by describing:* | 2.1 | **The type and quality of the data.** The type, quality, and relevance of the data and why these data sources are sufficient and suitable for capturing the target.<br>*Main reasons:* Enabling evaluation of the appropriateness and the validity and reliability of the data to be assessed[28,29]. |
| | 2.2 | **The data triangulation.** Whether and why the data come from different methodological approaches and the degree to which these approaches complement each other.<br>*Main reasons:* Using different and independent sources of information may complement each other in forming a more comprehensive assessment[24–27]. Enable evaluation of the data and potential mitigation of |

specific biases associated with certain methods.

2.3 **The data presentation**. How the data were structured and presented to the experts for their assessments and why.

*Main reasons:* Enable evaluation of the difficulty of the assessment task as well as how potential biases were mitigated, and limited cognitive resources were addressed[30,31].

2.4 **The access to the index measure**. For an assessment accuracy study, the extent the experts had access to the index measure and why (i.e., an assessment that is being compared to the best-estimate assessment), and how its information was weighted in their assessment.

*Main reasons:* Enable evaluation of any confirmation/verification biases and the validity/fairness of the comparison between the assessment and the index measure[3,46].

---

Evaluation – experts, materials and procedures

*Report the evaluation experts, materials, and procedures, by describing:*

3.1 **The expert and panel characteristics.** The characteristics of the experts and the panel, as well as how these characteristics are relevant for assessing the target.

*Main reasons:* To document the level of expertise and to get insight into experts' potential biases[34–36].

3.2 **The number of experts and panels.** The total number of experts and panels and how many experts/panels were assessing each case and why.

*Main reasons:* Providing information on the structure and consistency of the assessment method and how well individual human biases/errors are addressed[35,47,48].

3.3 **Blindness and conflicts of interest.** Whether and to what extent the experts are blind to the research aims and/or have any conflicts of interest.

*Main reasons:* Providing information regarding experts' potential biases and ethics[1].

3.4 **Instructions and training.** The instructions, training, and/or preparation that the experts specifically received for this assessment task and why they did or did not receive this.

*Main reasons:* Provide a better understanding of the difficulty of the assessment task and whether any specific measures were taken to limit biases[34–36].

3.5 **The assessment procedure.** The procedure that the experts followed for their assessment.

*Main reasons:* To provide information regarding the structure of the assessment task in order to 1) keep the assessment procedure consistent across the individual assessments, 2) to limit experts' potential biases, and 3) to address limited cognitive resources[30,35].

3.6 **The assessment response format.** The response format used by the experts for their individual assessments, what it included, and how it was structured.

*Main reasons:* To provide information regarding the structure of the assessment task in order to 1) keep the assessment procedure consistent across the individual assessments, 2) to limit experts' potential biases, and 3) to address limited cognitive resources[30,35].

3.7 **The data combination method.** The method or guidelines for how the data should be weighted, judged, and combined by the individual experts to reach a conclusion in their individual assessment.

*Main reasons:* Making the demands of experts explicit (e.g., see [32]). More complex information complicates the aggregation of data to reach a reliable conclusion[30,31].

3.8 **Independent expert assessments.** Whether and how the experts first evaluated the data individually and made their first individual assessments independently.

*Main reasons:* Provide information on the assessment procedure to know whether it is possible to calculate inter-rater reliability and to understand any consensus processes[1], e.g. how groupthink or overinfluence of one particular group member is limited.

| | | |
|---|---|---|
| | 3.9 | **The inter-rater and inter-panel reliability.** The inter-rater/inter-panel reliability, how it was calculated and evaluated, or why it was not possible to calculate it.<br>*Main reasons:* Provide information about the reliability of the individual and panel assessments on which the final assessment is based[1,32,49]. |
| | 3.10 | **The solution to disagreements.** The approach for solving (any) disagreements between the individual expert assessments, the rationale for the chosen approach, and potential problems that may have occurred and how these were assessed.<br>*Main reasons:* Providing information about the evaluation procedure, i.e., how the (potentially contradicting) individual assessments were combined into the final assessment[1]. |
| Validity<br>*Report what was assessed and how well, by describing:* | 4.1 | **The assessment description.** Description of *what* the assessment actually is.<br>*Main reasons:* Make it easier to evaluate the assessment. |
| | 4.2 | **The validity and standard.** Reflect on the degree of validity, and describe the standard that the method aims to achieve, *how* well the assessment method measures up to that degree, and how it compares with current standards.<br>*Main reasons:* Since the best-estimate assessment by definition is not the absolute truth, is highly dependent on the assessment method, and because it can be challenged and superseded by other methods, we consider it best conceptualized as a degree[39,40]. Reporting the degree is important for facilitating the evaluation of the standard status of an assessment, limiting overstating the achieved assessment, as well as simplifying comparisons with other (best-estimate) assessments. It will also help in avoiding having different assessments referred to as the "best"-estimate assessment when in fact being very different in quality. |

**Table S5 | *Examples of terms describing different assessment standards***

| Standard | Description |
|---|---|
| *Gold standard* | An error-free assessment |
| *Best-estimate assessment standard* | The best, practically feasible reference standard: It involves the state-of-the-art (i.e., the most valid and reliable) assessment method. |
| *New best-estimate assessment standard* | A new assessment method that provides an improved state-of-the-art (i.e., supersedes a previous best-estimate assessment method). |
| *Accepted reference/ criterion standard* | An assessment method that is not as accurate/comprehensive as the best-estimate assessment method but which provides sufficiently similar results to be acceptable to use in certain situations. |
| *Unrestricted best-estimate assessment standard* | When there are no restrictions for the assessment method (e.g., autopsy as compared to fMRI in identifying brain tumors). It can lead to procedures that are unreasonably comprehensive, complex, or unethical. |

*Note.* Different fields define/use these terms differently – here, we emphasize the importance of describing how it is being used within a specific project.

### Reports of the standards in 2022

The first author (V.E.) reviewed thirty randomly selected articles applying the assessment methods in 2022 and 2023 (i.e., five from each method from each year) and rated which standards were reported and which were lacking. Each reporting standard was rated using four categories: standard *not reported* (red); standard *reported vaguely or insufficiently* (orange); standard *(minimally) sufficiently reported* (green); or standard *not applicable for the study* (gray). Six randomly selected studies (i.e., one of each design from each year: studies 5, 6, 15, 20, 22, and 26 from Table 2) were reviewed by the second author (K.K.) to get insight into the accuracy of the ratings of the main author who reviewed the thirty studies. Most disagreements were between red/green and orange (*reported insufficiently*); not between green (*reported sufficiently*) and red (*not reported*). Many reporting standards were rather hard to find since they were often not explicitly described or reported scattered across an article. The authors disagreed on the reporting of 40 reporting standards across the six studies (33%) and discussed their disagreements to reach consensus.

They concluded the main rater (V.E.) potentially rated 23 reporting standards incorrectly (19%): Four standards were changed from orange to red (*3.9 The inter-rater and inter-panel reliability* of study 5; *1.3 History or lifetime information* of study 6; *2.3 The data presentation* of study 6; and *3.6 The assessment response format* of study 6). Two standards were changed from red to orange (*3.6 The assessment response format* of study 5 and *2.3 The data presentation* of study 26). Four standards were changed from orange to green (*1.4 The targeted time points* of study 15; *3.6 The assessment response format* of study 20; *3.6 The assessment response format* of study 26; and *3.8 Independent expert assessments* of study 26). Thirteen standards had to be changed from green to orange (*2.4 The access to index measure* of study 6; *3.4 Instructions and training* of study 6; *3.8 Independent expert assessments* of study 6; *4.2 The validity and standard* of study 6; *1.2 The number of time points* of study 20; *1.3 History or lifetime information* of study 20; *2.1 The type and quality of data* of study 20; *4.1 The assessment description* of study 20; *2.1 The type and quality of the data* of study 22; *3.1 The expert and panel characteristics* of study 22; and *3.3 Blindness and conflicts of interest* of study 22; *1.3 History or lifetime information* of study 26; and *3.1 The expert and panel characteristics* of study 26).

They concluded the second author (K.K.) rated eight reporting standards incorrectly (14%): Six standards were changed from orange into red (*3.4 Instructions and training* of study 6; *3.1 The expert and panel characteristics* of study 5; *3.3 Blindness and conflicts of interest* of study 15; *4.2 The validity and standard* of study 20; *1.4 The targeted time point(s)* of study 26; and *4.2 The validity and standard* of study 26). Two standards were changed from red to orange; (*4.1 The validity and standard* of study 22 and *3.9 The inter-rater and inter-panel reliability* of study 26). Two standards were changed from orange to green (*1.2 The number of time points* of study 6 and *1.1 The time period* of study 22). Four standards were changed from green to orange (*2.1 The type and quality of data* of study 6; *1.1 The time period* of study 20; *1.4 The targeted time points* of study 22; and *2.3 The data presentation* of study 26). One standard (*2.4 The access to the index measure* of study 22) was changed from orange to grey; one standard (*1.3 History or lifetime information* of study 15) was changed from grey to orange; and one standard (*3.9 The inter-rater and inter-panel reliability* of study 22) was changed from red to grey.

**References supplementary**

1. Bertens LCM, Broekhuizen BDL, Naaktgeboren CA, Rutten FH, Hoes AW, van Mourik Y, et al. Use of Expert Panels to Define the Reference Standard in Diagnostic Research: A Systematic Review of Published Methods and Reporting. PLoS Med. 2013 Oct 15;10(10):e1001531.
2. von Elm E, Altman DG, Egger M, Pocock SJ, Gøtzsche PC, Vandenbroucke JP. The Strengthening the Reporting of Observational Studies in Epidemiology (STROBE) Statement: Guidelines for reporting observational studies. Int J Surg. 2014 Dec 1;12(12):1495–9.
3. STARD Group, Bossuyt PM, Reitsma JB, Bruns DE, Gatsonis CA, Glasziou PP, et al. STARD 2015 : an updated list of essential items for reporting diagnostic accuracy studies. BMJ. 2015 Oct 28;351.
4. Moher D, Schulz KF, Simera I, Altman DG. Guidance for developers of health research reporting guidelines. PLoS Med. 2010 Feb 16;7(2):e1000217.
5. Handels RLH, Wolfs CAG, Aalten P, Bossuyt PMM, Joore MA, Leentjens AFG, et al. Optimizing the use of expert panel reference diagnoses in diagnostic studies of multidimensional syndromes. BMC Neurol. 2014 Oct 4;14:190.
6. Collins GS, Reitsma JB, Altman DG, Moons KGM. Transparent reporting of a multivariable prediction model for individual prognosis or diagnosis (TRIPOD): the TRIPOD statement. BMJ. 2015 Jan 7;350:g7594.
7. Schulz KF, Altman DG, Moher D, CONSORT Group. CONSORT 2010 statement: updated guidelines for reporting parallel group randomised trials. BMJ. 2010 Mar 23;340:c332.
8. Hoffmann TC, Glasziou PP, Boutron I, Milne R, Perera R, Moher D, et al. Better reporting of interventions: template for intervention description and replication (TIDieR) checklist and guide. BMJ. 2014 Mar 7;348(mar07 3):g1687–g1687.
9. Jünger S, Payne SA, Brine J, Radbruch L, Brearley SG. Guidance on Conducting and REporting DElphi Studies (CREDES) in palliative care: Recommendations based on a methodological systematic review. Palliat Med. 2017 Sep;31(8):684–706.
10. Kirkham JJ, Gorst S, Altman DG, Blazeby JM, Clarke M, Devane D, et al. Core Outcome Set–STAndards for Reporting: The COS-STAR Statement. PLOS Med. 2016 okt;13(10):e1002148.
11. Vasey B, Nagendran M, Campbell B, Clifton DA, Collins GS, Denaxas S, et al. Reporting guideline for the early stage clinical evaluation of decision support systems driven by artificial intelligence: DECIDE-AI. BMJ. 2022 May 18;377:e070904.
12. Reitsma JB, Rutjes AWS, Khan KS, Coomarasamy A, Bossuyt PM. A review of solutions for diagnostic accuracy studies with an imperfect or missing reference standard. J Clin Epidemiol. 2009 Jan 1;62(8):797–806.
13. Husereau D, Drummond M, Augustovski F, de Bekker-Grob E, Briggs AH, Carswell C, et al. Consolidated Health Economic Evaluation Reporting Standards 2022 (CHEERS 2022) statement: updated reporting guidance for health economic evaluations. BMC Med. 2022 Jan 12;20(1):23.
14. Spranger J, Homberg A, Sonnberger M, Niederberger M. Reporting guidelines for Delphi techniques in health sciences: A methodological review. Z Evidenz Fortbild Qual Im Gesundheitswesen. 2022 Aug;172:1–11.
15. Robin W M Vernooij, Pablo Alonso-Coello, Melissa Brouwers, Laura Martínez García, CheckUp Panel. Reporting Items for Updated Clinical Guidelines: Checklist for the Reporting of Updated Guidelines (CheckUp). PLoS Med. 2017 Jan 1;14(1):e1002207–e1002207.
16. American Psychiatric Association. DSM-5 Task Force, American Psychiatric Association. Diagnostic and statistical manual of mental disorders : DSM-5. 5. ed. American Psychiatric Association; 2013.
17. Spitzer RL. Psychiatric diagnosis: Are clinicians still necessary? Compr Psychiatry. 1983 Sep;24(5):399–411.
18. Ben-Zeev D, Young MA. Accuracy of hospitalized depressed patients' and healthy controls' retrospective symptom reports: an experience sampling study. J Nerv Ment Dis. 2010 Apr;198(4):280–5.

19. Bradburn NM, Rips LJ, Shevell SK. Answering Autobiographical Questions: The Impact of Memory and Inference on Surveys. Science. 1987 Apr 10;236(4798):157–61.
20. Caruana EJ, Roman M, Hernández-Sánchez J, Solli P. Longitudinal studies. J Thorac Dis. 2015 Nov;7(11):E537–40.
21. Stone AA, Shiffman S. Ecological Momentary Assessment (Ema) in Behavioral Medicine. Ann Behav Med. 1994 Jan 1;16(3):199–202.
22. Kim J, Marcusson-Clavertz D, Yoshiuchi K, Smyth JM. Potential benefits of integrating ecological momentary assessment data into mHealth care systems. Biopsychosoc Med. 2019;13:19.
23. Shiffman S, Stone AA, Hufford MR. Ecological momentary assessment. Annu Rev Clin Psychol. 2008 Jan 1;4:1–32.
24. Mathison S. Why Triangulate? Educ Res. 1988 Mar 1;17(2):13–7.
25. Lawlor D a., Tilling K, Smith G d. Triangulation in aetiological epidemiology. Int J Epidemiol. 2016 Dec 1;45(6):1866–86.
26. Lahat D, Adali T, Jutten C. Multimodal Data Fusion: An Overview of Methods, Challenges, and Prospects. Proc IEEE Proc IEEE. 2015 Sep 1;103(9):1449–77.
27. Wang L, Wu J, Huang SL, Zheng L, Xu X, Zhang L, et al. An Efficient Approach to Informative Feature Extraction from Multimodal Data. 2018;
28. Cronbach LJ, Meehl PE. Construct validity in psychological tests. Psychol Bull. 1955 Jul;52(4):281–302.
29. Scott O Lilienfeld, Katheryn eSauvigne, Steven Jay Lynn, Robert D Latzman, Robin eCautin, Irwin D. Waldman. Fifty Psychological and Psychiatric Terms to Avoid: A List of Inaccurate, Misleading, Misused, Ambiguous, and Logically Confused Words and Phrases. Front Psychol. 2015 Aug 1;6.
30. Grove WM, Meehl PE. Comparative Efficiency of Informal (Subjective, Impressionistic) and Formal (Mechanical, Algorithmic) Prediction Procedures: The Clinical-Statistical Controversy. Psychol Public Policy Law. 1996 Jun 1;2(2):293–323.
31. Meehl PE. Clinical versus statistical prediction : a theoretical analysis and a review of the evidence. University of Minnesota Press; 1973.
32. Klein DN, Ouimette PC, Kelly HS, Ferro T, Riso LP. Test-retest reliability of team consensus best-estimate diagnoses of axis I and II disorders in a family study. Am J Psychiatry. 1994 Jul;151(7):1043–7.
33. Miller P r., Dasher R, Collins R, Griffiths P, Brown F. Inpatient diagnostic assessments: 1. Accuracy of structured vs. Unstructured interviews. Psychiatry Res. 2001 Dec 31;105(3):255–64.
34. Bowes S m., Ammirati R j., Costello T h., Basterfield C, Lilienfeld S o. Cognitive biases, heuristics, and logical fallacies in clinical practice: A brief field guide for practicing clinicians and supervisors. Prof Psychol Res Pract. 2020 Jan 1;51(5):435–45.
35. Saposnik G, Redelmeier D, Ruff CC, Tobler PN. Cognitive biases associated with medical decisions: a systematic review. BMC Med Inform Decis Mak. 2016 Nov 3;16:1–14.
36. Faust D. Research on Human Judgment and Its Application to Clinical Practice. Prof Psychol Res Pract. 1986 Oct 1;17(5):420–30.
37. Claassen JAHR. The Gold Standard: Not A Golden Standard. BMJ. 2005 May 14;330(7500):1121–1121.
38. Versi E. ‘Gold Standard’ Is An Appropriate Term. BMJ. 1992 Jul 18;305(6846):187–187.
39. American Educational Research Association, American Psychological Association, National Council on Measurement in Education. Standards for educational and psychological testing. Washington, DC: American Educational Research Association; 2014. 230 p.
40. Popper K. Objective knowledge : an evolutionary approach. Rev. ed. Clarendon Press; 1979.
41. Gearing RE, Mian IA, Barber J, Ickowicz A. A methodology for conducting retrospective chart review research in child and adolescent psychiatry. J Can Acad Child Adolesc Psychiatry J Acad Can Psychiatr Enfant Adolesc. 2006 Aug;15(3):126–34.
42. Jivraj S, Goodman A, Ploubidis GB, de Oliveira C. Testing Comparability Between Retrospective Life History Data and Prospective Birth Cohort Study Data. J Gerontol B Psychol Sci Soc Sci. 2020

Jan 1;75(1):207–17.
43. Worster A, Haines T. Advanced statistics: understanding medical record review (MRR) studies. Acad Emerg Med Off J Soc Acad Emerg Med. 2004 Feb;11(2):187–92.
44. Odgers CL, Mulvey EP, Skeem JL, Gardner W, Lidz CW, Schubert C. Capturing the ebb and flow of psychiatric symptoms with dynamical systems models. Am J Psychiatry. 2009 May;166(5):575–82.
45. Schoevers RA, van Borkulo CD, Lamers F, Servaas MN, Bastiaansen JA, Beekman ATF, et al. Affect fluctuations examined with ecological momentary assessment in patients with current or remitted depression and anxiety disorders. Psychol Med. 2021 Aug;51(11):1906–15.
46. Kranzler HR, Tennen H, Babor TF, Kadden RM, Rounsaville BJ. Validity of the longitudinal, expert, all data procedure for psychiatric diagnosis in patients with psychoactive substance use disorders. Drug Alcohol Depend. 1997 Apr 14;45(1–2):93–104.
47. Gagnon R, Charlin B, Coletti M, Sauvé E, van der Vleuten C. Assessment in the context of uncertainty: how many members are needed on the panel of reference of a script concordance test? Med Educ. 2005 Mar;39(3):284–91.
48. van Houten CB, Naaktgeboren CA, Ashkenazi-Hoffnung L, Ashkenazi S, Avis W, Chistyakov I, et al. Expert panel diagnosis demonstrated high reproducibility as reference standard in infectious diseases. J Clin Epidemiol. 2019 Aug;112:20–7.
49. McHugh ML. Interrater reliability: the kappa statistic. Biochem Medica. 2012;22(3):276–82.

**Delphi references**

1. Abong J, Dalay V, Langley I, Tomeny E, Marcelo D, Mendoza V, et al. Use of GeneXpert and the role of an expert panel in improving clinical diagnosis of smear-negative tuberculosis cases. Quinn F, editor. PLOS ONE. 2019 Dec 30;14(12):e0227093.
2. Allison KH, Reisch LM, Carney PA, Weaver DL, Schnitt SJ, O'Malley FP, et al. Understanding diagnostic variability in breast pathology: Lessons learned from an expert consensus review panel. Histopathology. 2014 Aug;65(2):240–51.
3. Alway Y, Gould KR, Johnston L, McKenzie D, Ponsford J. A prospective examination of Axis I psychiatric disorders in the first 5 years following moderate to severe traumatic brain injury. Psychol Med. 2016 Apr;46(6):1331–41.
4. Andersson M, Bäckström M, Ivarsson T, Råstam M, Jarbin H. Validity of the Brief Child and Family Phone Interview by comparison with Longitudinal Expert All Data diagnoses in outpatients. Scand J Child Adolesc Psychiatry Psychol. 2020 Oct 18;6(2):83–90.
5. Aydin S, Siebelink BM, Crone MR, van Ginkel JR, Numans ME, Vermeiren RRJM, et al. The diagnostic process from primary care to child and adolescent mental healthcare services: the incremental value of information conveyed through referral letters, screening questionnaires and structured multi-informant assessment. BJPsych Open. 2022 Apr 7;8(3):e81.
6. Baker MW, Whitney JD, Lowe JR, Liao S, Zimmerman D, Mosqueda L. Full-Thickness and Unstageable Pressure Injuries That Develop in Nursing Home Residents Despite Consistently Good Quality Care. J Wound Ostomy Cont Nurs Off Publ Wound Ostomy Cont Nurses Soc. 2016;43(5):464–70.
7. Bech P, Timmerby N, Martiny K, Lunde M, Soendergaard S. Psychometric evaluation of the Major Depression Inventory (MDI) as depression severity scale using the LEAD (Longitudinal Expert Assessment of All Data) as index of validity. BMC Psychiatry. 2015 Aug 5;15:190.
8. Beeney JE, Wright AGC, Stepp SD, Hallquist MN, Lazarus SA, Beeney JRS, et al. Disorganized attachment and personality functioning in adults: A latent class analysis. Personal Disord Theory Res Treat. 2017;8:206–16.
9. Berghuis H, Bandell CC, Krueger RF. Predicting dropout using DSM-5 Section II personality disorders, and DSM-5 Section III personality traits, in a (day)clinical sample of personality disorders. Personal Disord. 2021 Jul;12(4):331–8.
10. Black DW, Coryell WH, Crowe RR, McCormick B, Shaw MC, Allen J. A direct, controlled, blind

family study of DSM-IV pathological gambling. J Clin Psychiatry. 2014 Mar;75(3):215–21.
11. Blackmore R, Gray KM, Melvin GA, Newman L, Boyle JA, Gibson-Helm M. Identifying post-traumatic stress disorder in women of refugee background at a public antenatal clinic. Arch Womens Ment Health. 2022 Feb;25(1):191–8.
12. Bone D, Bishop S, Gupta R, Lee S, Narayanan SS. Acoustic-Prosodic and Turn-Taking Features in Interactions with Children with Neurodevelopmental Disorders. In: Interspeech 2016 [Internet]. ISCA; 2016 [cited 2023 May 14]. p. 1185–9. Available from: https://www.isca-speech.org/archive/interspeech_2016/bone16_interspeech.html
13. Bradshaw J, Shi D, Hendrix CL, Saulnier C, Klaiman C. Neonatal neurobehavior in infants with autism spectrum disorder. Dev Med Child Neurol. 2022 May;64(5):600–7.
14. Bertens LCM, van Mourik Y, Rutten FH, Cramer MJM, Lammers JWJ, Hoes AW, et al. Staged decision making was an attractive alternative to a plenary approach in panel diagnosis as reference standard. J Clin Epidemiol. 2015 Apr 1;68(4):418–25.
15. Breivik R, Wilberg T, Evensen J, Røssberg JI, Dahl HSJ, Pedersen G. Countertransference feelings and personality disorders: a psychometric evaluation of a brief version of the Feeling Word Checklist (FWC-BV). BMC Psychiatry. 2020 Mar 30;20(1):141.
16. Brian J, Bryson SE, Smith IM, Roberts W, Roncadin C, Szatmari P, et al. Stability and change in autism spectrum disorder diagnosis from age 3 to middle childhood in a high-risk sibling cohort. Autism. 2016 Oct 1;20(7):888–92.
17. Brunyé TT, Mercan E, Weaver DL, Elmore JG. Accuracy is in the eyes of the pathologist: The visual interpretive process and diagnostic accuracy with digital whole slide images. J Biomed Inform. 2017 Feb;66:171–9.
18. Bunte TL, Laschen S, Schoemaker K, Hessen DJ, van der Heijden PGM, Matthys W. Clinical usefulness of observational assessment in the diagnosis of DBD and ADHD in preschoolers. J Clin Child Adolesc Psychol Off J Soc Clin Child Adolesc Psychol Am Psychol Assoc Div 53. 2013;42(6):749–61.
19. Bøen E, Hummelen B, Elvsåshagen T, Boye B, Andersson S, Karterud S, et al. Different impulsivity profiles in borderline personality disorder and bipolar II disorder. J Affect Disord. 2015 Jan 1;170:104–11.
20. Campbell DJ, Shic F, Macari S, Chawarska K. Gaze response to dyadic bids at 2 years related to outcomes at 3 years in autism spectrum disorders: a subtyping analysis. J Autism Dev Disord. 2014 Feb;44(2):431–42.
21. Chawarska K, Macari S, Shic F. Decreased spontaneous attention to social scenes in 6-month-old infants later diagnosed with autism spectrum disorders. Biol Psychiatry. 2013 Aug 1;74(3):195–203.
22. Coccaro EF, Berman ME, McCloskey MS. Development of a screening questionnaire for DSM-5 intermittent explosive disorder (IED-SQ). Compr Psychiatry. 2017 Apr;74:21–6.
23. Colvert E, Tick B, McEwen F, Stewart C, Curran SR, Woodhouse E, et al. Heritability of Autism Spectrum Disorder in a UK Population-Based Twin Sample. JAMA Psychiatry. 2015 May;72(5):415–23.
24. Cowan KJ, Tandias A, Arndt B, Hanrahan L, Mundt M, Guilbert TW. Defining Asthma: Validating Automated Electronic Health Record Algorithm With Expert Panel Diagnosis. In: A104 ASTHMA EPIDEMIOLOGY [Internet]. American Thoracic Society; 2014 [cited 2023 May 12]. p. A2297–A2297. (American Thoracic Society International Conference Abstracts). Available from: https://www.atsjournals.org/doi/10.1164/ajrccm-conference.2014.189.1_MeetingAbstracts.A2297
25. Cumming K, Hoyle GE, Hutchison JD, Soiza RL. Prevalence, incidence and etiology of hyponatremia in elderly patients with fragility fractures. PloS One. 2014;9(2):e88272.
26. Davis KAS, Bashford O, Jewell A, Shetty H, Stewart RJ, Sudlow CLM, et al. Using data linkage to electronic patient records to assess the validity of selected mental health diagnoses in English Hospital Episode Statistics (HES). PLOS ONE. 2018 mrt;13(3):e0195002.
27. Deckers A, Muris P, Roelofs J. Being on Your Own or Feeling Lonely? Loneliness and Other Social Variables in Youths with Autism Spectrum Disorders. Child Psychiatry Hum Dev. 2017

Oct;48(5):828–39.
28. Dereboy F, Dereboy Ç, Eskin M. Validation of the DSM–5 Alternative Model Personality Disorder Diagnoses in Turkey, Part 1: LEAD Validity and Reliability of the Personality Functioning Ratings. J Pers Assess. 2018 Nov 2;100(6):603–11.
29. Di Florio A, Forty L, Gordon-Smith K, Heron J, Jones L, Craddock N, et al. Perinatal episodes across the mood disorder spectrum. JAMA Psychiatry. 2013 Feb;70(2):168–75.
30. Duffy A, Horrocks J, Doucette S, Keown-Stoneman C, McCloskey S, Grof P. Childhood anxiety: an early predictor of mood disorders in offspring of bipolar parents. J Affect Disord. 2013 Sep 5;150(2):363–9.
31. Edmonds EC, Smirnov DS, Thomas KR, Graves LV, Bangen KJ, Delano-Wood L, et al. Data-Driven vs Consensus Diagnosis of MCI: Enhanced Sensitivity for Detection of Clinical, Biomarker, and Neuropathologic Outcomes. Neurology. 2021 Sep 28;97(13):e1288–99.
32. Elias R, Lord C. Diagnostic stability in individuals with autism spectrum disorder: insights from a longitudinal follow-up study. J Child Psychol Psychiatry. 2022 Sep;63(9):973–83.
33. Elmore JG, Nelson HD, Pepe MS, Longton GM, Tosteson ANA, Geller B, et al. Variability in Pathologists' Interpretations of Individual Breast Biopsy Slides: A Population Perspective. Ann Intern Med. 2016 May 17;164(10):649–55.
34. Ferreira-Maia AP, Boronat AC, Boarati MA, Fu-I L, Wang YP. Evaluation of Bipolar Disorder in Children and Adolescents Referred to a Mood Service: Diagnostic Pathways and Manic Dimensions. J Psychiatr Pract. 2016 Nov;22(6):429–41.
35. Fiedorowicz JG, Jancic D, Potash JB, Butcher B, Coryell WH. Vascular Mortality in Participants of a Bipolar Genomics Study. Psychosomatics. 2014 Sep 1;55(5):485–90.
36. Folmo EJ, Stänicke E, Johansen MS, Pedersen G, Kvarstein EH. Development of therapeutic alliance in mentalization-based treatment-Goals, Bonds, and Tasks in a specialized treatment for borderline personality disorder. Psychother Res J Soc Psychother Res. 2021 Jun;31(5):604–18.
37. Gangi DN, Ibañez LV, Messinger DS. Joint attention initiation with and without positive affect: risk group differences and associations with ASD symptoms. J Autism Dev Disord. 2014 Jun;44(6):1414–24.
38. Gao R, Zhao S, Aishanjiang K, Cai H, Wei T, Zhang Y, et al. Deep learning for differential diagnosis of malignant hepatic tumors based on multi-phase contrast-enhanced CT and clinical data. J Hematol OncolJ Hematol Oncol. 2021 Sep 26;14:154.
39. Garner BR, Scott CK, Dennis ML, Funk RR. The relationship between recovery and health-related quality of life. J Subst Abuse Treat. 2014 Oct;47(4):293–8.
40. Girault JB, Swanson MR, Meera SS, Grzadzinski RL, Shen MD, Burrows CA, et al. Quantitative trait variation in ASD probands and toddler sibling outcomes at 24 months. J Neurodev Disord. 2020 Feb 5;12(1):5.
41. Gurriarán X, Rodríguez-López J, Flórez G, Pereiro C, Fernández JM, Fariñas E, et al. Relationships between substance abuse/dependence and psychiatric disorders based on polygenic scores. Genes Brain Behav. 2019 Mar;18(3):e12504.
42. Handels RLH, Wolfs CAG, Aalten P, Bossuyt PMM, Joore MA, Leentjens AFG, et al. Optimizing the use of expert panel reference diagnoses in diagnostic studies of multidimensional syndromes. BMC Neurol. 2014 Oct 4;14:190.
43. Hendriks E, Muris P, Meesters C, Houben K. Childhood Disorder: Dysregulated Self-Conscious Emotions? Psychopathological Correlates of Implicit and Explicit Shame and Guilt in Clinical and Non-clinical Children and Adolescents. Front Psychol. 2022;13:822725.
44. Hernández-Chan GS, Ceh-Varela EE, Sanchez-Cervantes JL, Villanueva-Escalante M, Rodríguez-González A, Pérez-Gallardo Y. Collective intelligence in medical diagnosis systems: A case study. Comput Biol Med. 2016 Jul 1;74:45–53.
45. Hofvander B, Anckarsäter H, Wallinius M, Billstedt E. Mental health among young adults in prison: the importance of childhood-onset conduct disorder. BJPsych Open. 2017 Mar;3(2):78–84.
46. Högberg C, Billstedt E, Björck C, Björck PO, Ehlers S, Gustle LH, et al. Diagnostic validity of the

MINI-KID disorder classifications in specialized child and adolescent psychiatric outpatient clinics in Sweden. BMC Psychiatry. 2019 May 9;19(1):142.
47. Hummelen B, Pedersen G, Wilberg T, Karterud S. Poor Validity of the DSM-IV Schizoid Personality Disorder Construct as a Diagnostic Category. J Personal Disord. 2015 Jun;29(3):334–46.
48. Jarbin H, Ivarsson T, Andersson M, Bergman H, Skarphedinsson G. Screening efficiency of the Mood and Feelings Questionnaire (MFQ) and Short Mood and Feelings Questionnaire (SMFQ) in Swedish help seeking outpatients. Montazeri A, editor. PLOS ONE. 2020 Mar 25;15(3):e0230623.
49. Jin H, Chien S, Meijer E, Khobragade P, Lee J. Learning From Clinical Consensus Diagnosis in India to Facilitate Automatic Classification of Dementia: Machine Learning Study. JMIR Ment Health. 2021 May 10;8(5):e27113.
50. Katzin S, Andiné P, Hofvander B, Billstedt E, Wallinius M. Exploring Traumatic Brain Injuries and Aggressive Antisocial Behaviors in Young Male Violent Offenders. Front Psychiatry. 2020;11:507196.
51. Kim SH, Bal VH, Benrey N, Choi YB, Guthrie W, Colombi C, et al. Variability in Autism Symptom Trajectories Using Repeated Observations From 14 to 36 Months of Age. J Am Acad Child Adolesc Psychiatry. 2018 Nov;57(11):837-848.e2.
52. Kuhn E, Du X, McGrath K, Coveney S, O'Regan N, Richardson S, et al. Validation of a consensus method for identifying delirium from hospital records. PloS One. 2014;9(11):e111823.
53. Kuru Y, Nishiyama T, Sumi S, Suzuki F, Shiino T, Kimura T, et al. Practical applications of brief screening questionnaires for autism spectrum disorder in a psychiatry outpatient setting. Int J Methods Psychiatr Res. 2020 Nov 20;30(2):e1857.
54. Lamers F, Cui L, Hickie IB, Roca C, Machado-Vieira R, Zarate CA, et al. Familial aggregation and heritability of the melancholic and atypical subtypes of depression. J Affect Disord. 2016 Nov 1;204:241–6.
55. Lewis KC, Meehan KB, Cain NM, Wong PS, Clemence AJ, Stevens J, et al. Impairments in Object Relations and Chronicity of Suicidal Behavior in Individuals With Borderline Personality Disorder. J Personal Disord. 2016 Feb;30(1):19–34.
56. Mataix-Cols D, Billotti D, Fernández de la Cruz L, Nordsletten AE. The London field trial for hoarding disorder. Psychol Med. 2013 Apr;43(4):837–47.
57. McKenzie DP, Downing MG, Ponsford JL. Key Hospital Anxiety and Depression Scale (HADS) items associated with DSM-IV depressive and anxiety disorder 12-months post traumatic brain injury. J Affect Disord. 2018 Aug 15;236:164–71.
58. Molina TJ, Bluthgen MV, Chalabreysse L, Montpréville VT de, Muret A de, Dubois R, et al. Impact of expert pathologic review of thymic epithelial tumours on diagnosis and management in a real-life setting: A RYTHMIC study. Eur J Cancer. 2021 Jan 1;143:158–67.
59. Mooney MA, Bhatt P, Hermosillo RJM, Ryabinin P, Nikolas M, Faraone SV, et al. Smaller total brain volume but not subcortical structure volume related to common genetic risk for ADHD. Psychol Med. 2021 Jun;51(8):1279–88.
60. Morrison EH, Sorkin D, Mosqueda L, Ayutyanont N. Validity and Reliability of the Scale to Report Emotional Stress Signs–Multiple Sclerosis (STRESS-MS) in Assessing Abuse and Neglect of Adults With Multiple Sclerosis. Int J MS Care. 2022 Jan 1;24(1):18–24.
61. Muris P, Meesters C, Heijmans J, van Hulten S, Kaanen L, Oerlemans B, et al. Lack of guilt, guilt, and shame: a multi-informant study on the relations between self-conscious emotions and psychopathology in clinically referred children and adolescents. Eur Child Adolesc Psychiatry. 2016 Apr 1;25(4):383–96.
62. Nigg JT, Gustafsson HC, Karalunas SL, Ryabinin P, McWeeney SK, Faraone SV, et al. Working Memory and Vigilance as Multivariate Endophenotypes Related to Common Genetic Risk for Attention-Deficit/Hyperactivity Disorder. J Am Acad Child Adolesc Psychiatry. 2018 Mar;57(3):175–82.
63. Nishiyama T, Sumi S, Watanabe H, Suzuki F, Kuru Y, Shiino T, et al. The Kiddie Schedule for

Affective Disorders and Schizophrenia Present and Lifetime Version (K-SADS-PL) for DSM-5: A validation for neurodevelopmental disorders in Japanese outpatients. Compr Psychiatry. 2020 Jan 1;96:152148.
64. North CS, Simic Z, Burruss J. Design, Implementation, and Assessment of a Public Comprehensive Specialty Care Program for Early Psychosis. J Psychiatr Pract. 2019 Mar;25(2):91–102.
65. Nyrenius J, Eberhard J, Ghaziuddin M, Gillberg C, Billstedt E. Prevalence of Autism Spectrum Disorders in Adult Outpatient Psychiatry. J Autism Dev Disord. 2022 Sep 1;52(9):3769–79.
66. Osório FL, Loureiro SR, Hallak JEC, Machado-de-Sousa JP, Ushirohira JM, Baes CVW, et al. Clinical validity and intrarater and test-retest reliability of the Structured Clinical Interview for DSM-5 - Clinician Version (SCID-5-CV). Psychiatry Clin Neurosci. 2019 Dec;73(12):754–60.
67. Ousley O, Evans AN, Fernandez-Carriba S, Smearman EL, Rockers K, Morrier MJ, et al. Examining the Overlap between Autism Spectrum Disorder and 22q11.2 Deletion Syndrome. Int J Mol Sci. 2017 May 18;18(5):1071.
68. Pedersen G, Karterud S, Hummelen B, Wilberg T. The impact of extended longitudinal observation on the assessment of personality disorders. Personal Ment Health. 2013 Nov;7(4):277–87.
69. Peters ME, Rao V, Bechtold KT, Roy D, Sair HI, Leoutsakos JM, et al. Head injury serum markers for assessing response to trauma: Design of the HeadSMART study. Brain Inj. 2017;31(3):370–8.
70. Reas DL, Rø Ø, Karterud S, Hummelen B, Pedersen G. Eating disorders in a large clinical sample of men and women with personality disorders: Eating Disorders In Personality Disorders. Int J Eat Disord. 2013 Dec;46(8):801–9.
71. Yonashiro-Cho JMF, Gassoumis ZD, Wilber KH, Homeier DC. Improving forensics: Characterizing injuries among community-dwelling physically abused older adults. J Am Geriatr Soc. 2021 Aug;69(8):2252–61.
72. Houten CB van, Naaktgeboren CA, Ashkenazi-Hoffnung L, Ashkenazi S, Avis W, Chistyakov I, et al. Expert panel diagnosis demonstrated high reproducibility as reference standard in infectious diseases. J Clin Epidemiol. 2019 Aug 1;112:20–7.
73. Platts-Mills TF, Dayaa JA, Reeve BB, Krajick K, Mosqueda L, Haukoos JS, et al. Development of the Emergency Department Senior Abuse Identification (ED Senior AID) tool. J Elder Abuse Negl. 2018;30(4):247–70.
74. Reas DL, Pedersen G, Karterud S, Rø Ø. Self-harm and suicidal behavior in borderline personality disorder with and without bulimia nervosa. J Consult Clin Psychol. 2015 Jun;83(3):643–8.
75. Ridenour JM, Lewis KC, Poston JM, Ciecalone DN. Performance-based assessment of social cognition in borderline versus psychotic psychopathology. Rorschachiana. 2019;40:95–111.
76. Ridenour JM, Lewis KC, Siefert CJ, Pitman SR, Knauss D, Stein MB. Card pull effects of the Thematic Apperception Test using the Social Cognition and Object Relations-Global Rating Method on complex psychiatric sample. Clin Psychol Psychother. 2021 Sep;28(5):1079–90.
77. Rodríguez-López J, Flórez G, Blanco V, Pereiro C, Fernández JM, Fariñas E, et al. Genome wide analysis of rare copy number variations in alcohol abuse or dependence. J Psychiatr Res. 2018 Aug 1;103:212–8.
78. Platts-Mills TF, Encarnacion JA, Bin Shams R, Hurka-Richardson K, Rosen T, Cannell B. Reliability of the longitudinal experts all data (LEAD) methodology for determining the presence of elder mistreatment. J Elder Abuse Negl. 2021 Oct 20;33(5):385–97.
79. Snyder SM, Rugino TA, Hornig M, Stein MA. Integration of an EEG biomarker with a clinician's ADHD evaluation. Brain Behav. 2015 Apr;5(4):e00330.
80. Steffenburg H, Steffenburg S, Gillberg C, Billstedt E. Children with autism spectrum disorders and selective mutism. Neuropsychiatr Dis Treat. 2018 May 7;14:1163–9.
81. Teeters DA, Moua T, Li G, Kashyap R, Biehl M, Kaur R, et al. Mild Cognitive Impairment and Risk of Critical Illness. Crit Care Med. 2016 Nov;44(11):2045–51.
82. ter Huurne ED, de Haan HA, ten Napel-Schutz MC, Postel MG, Menting J, van der Palen J, et al. Is the Eating Disorder Questionnaire-Online (EDQ-O) a valid diagnostic instrument for the DSM-IV-TR classification of eating disorders? Compr Psychiatry. 2015 Feb 1;57:167–76.

**Standard reports in 2022 references**

1. Ahuja K, Kandwal P, Ifthekar S, Sudhakar PV, Nene A, Basu S, et al. Development of Tuberculosis Spine Instability Score (TSIS): An evidence-based and expert consensus-based content validation study among spine surgeons. Spine. 2022;47(3):242-51.
2. Asbjornsdottir B, Lauth B, Fasano A, Thorsdottir I, Karlsdottir I, Gudmundsson LS, et al. Meals, Microbiota and Mental Health in Children and Adolescents (MMM-Study): A protocol for an observational longitudinal case-control study. PLoS One. 2022;17(9)
3. Aydin S, Siebelink BM, Crone MR, van Ginkel JR, Numans ME, Vermeiren RR, et al. The diagnostic process from primary care to child and adolescent mental healthcare services: the incremental value of information conveyed through referral letters, screening questionnaires and structured multi-informant assessment. BJPsych Open. 2022;8(3)
4. Berthelot N, Garon-Bissonnette J, Jomphe V, Doucet-Beaupré H, Bureau A, Maziade M. Childhood trauma may increase risk of psychosis and mood disorder in genetically high-risk children and adolescents by enhancing the accumulation of risk indicators. Schizophr Bull Open. 2022;3(1)
5. Blackmore R, Gray KM, Melvin GA, Newman L, Boyle JA, Gibson-Helm M. Identifying post-traumatic stress disorder in women of refugee background at a public antenatal clinic. Arch Womens Ment Health. 2022;25(1):191-8.
6. Bradshaw J, Shi D, Hendrix CL, Saulnier C, Klaiman C. Neonatal neurobehavior in infants with autism spectrum disorder. Dev Med Child Neurol. 2022;64(5):600-7.
7. Bulten W, Kartasalo K, Chen PHC, Ström P, Pinckaers H, Nagpal K, et al. Artificial intelligence for diagnosis and Gleason grading of prostate cancer: the PANDA challenge. Nat Med. 2022;28(1):154-63.
8. Campbell JP, Chiang MF, Chen JS, Moshfeghi DM, Nudleman E, Ruambivoonsuk P, et al. Artificial intelligence for retinopathy of prematurity: validation of a vascular severity scale against international expert diagnosis. Ophthalmology. 2022;129(7)
9. Dow D, Holbrook A, Toolan C, McDonald N, Sterrett K, Rosen N, et al. The brief observation of symptoms of autism (BOSA): development of a new adapted assessment measure for remote telehealth administration through COVID-19 and beyond. J Autism Dev Disord. 2022;52(12):5383-94.
10. Du Y, Bara M, Katlariwala P, Croutze R, Resch K, Porter J, et al. Effect of training on resident inter-reader agreement with American College of Radiology Thyroid Imaging Reporting and Data System. World J Radiol. 2022;14(1):19.
11. Estrada-Jaramillo S, Quintero-Cadavid CP, Andrade-Carrillo R, Gómez-Cano S, Erazo-Osorio JJ, Zapata-Ospina JP, et al. Do Children of Patients with Bipolar Disorder have a Worse Perception of

Sleep Quality? Rev Colomb Psiquiatr (Engl Ed). 2022;51(1):25-34.
12. Gunaratna GP, Mohammad SS, Blyth CC, Clark J, Crawford N, Marshall H, et al. Postinfectious Acute Cerebellar Syndromes in Children: A Nationally Ascertained Case Series From Australia 2013–2018. J Child Neurol. 2022;37(7):617-23.
13. Hagens LA, Van der Ven FL, Heijnen NF, Smit MR, Gietema HA, Gerretsen SC, et al. Improvement of an interobserver agreement of ARDS diagnosis by adding additional imaging and a confidence scale. Front Med. 2022;9:950827.
14. Hendriks E, Muris P, Meesters C, Houben K. Childhood disorder: dysregulated self-conscious emotions? Psychopathological correlates of implicit and explicit shame and guilt in clinical and non-clinical children and adolescents. Front Psychol. 2022;13:822725.
15. Hesam-Shariati S, Overs BJ, Roberts G, Toma C, Watkeys OJ, Green MJ, et al. Epigenetic signatures relating to disease-associated genotypic burden in familial risk of bipolar disorder. Transl Psychiatry. 2022;12(1):310.
16. Khan AM, Ahmed S, Chowdhury NH, Islam MS, McCollum ED, King C, et al. Developing a video expert panel as a reference standard to evaluate respiratory rate counting in paediatric pneumonia diagnosis: protocol for a cross-sectional study. BMJ Open. 2022;12(11)
17. Kim SY, Oh M, Bong G, Song DY, Yoon NH, Kim JH, et al. Diagnostic validity of Autism Diagnostic Observation schedule, (K-ADOS-2) in the Korean population. Mol Autism. 2022;13(1):30.
18. Landa RJ, Reetzke R, Holingue CB, Herman D, Hess CR. Diagnostic Stability and Phenotypic Differences Among School-Age Children Diagnosed With ASD Before Age 2. Front Psychiatry. 2022;13:805686.
19. Leroux A, Frey KP, Crainiceanu CM, Obremskey WT, Stinner DJ, Bosse MJ, et al. Defining Incidence of Acute Compartment Syndrome in the Research Setting: A Proposed Method From the PACS Study. J Orthop Trauma. 2022;36
20. Lindhardt L, Nilsson LS, Munk-Jørgensen P, Mortensen OS, Simonsen E, Nordgaard J. Unrecognized schizophrenia spectrum and other mental disorders in youth disconnected from education and work-life. Front Psychiatry. 2022;13:1015616.
21. Loots FJ, Smits M, Hopstaken RM, Jenniskens K, Schroeten FH, Van den Bruel A, et al. New clinical prediction model for early recognition of sepsis in adult primary care patients: a prospective diagnostic cohort study of development and external validation. Br J Gen Pract. 2022;72(719)
22. Mackenhauer J, Winsløv JH, Holmskov J, Brødsgaard I, Larsen TG, Mainz J. Analysis of suicides reported as adverse events in psychiatry resulted in nine quality improvement initiatives. Crisis. 2022;43(4):307-14.
23. Maziade M, Bureau A, Jomphe V, Gagné AM. Retinal function and preclinical risk traits in children and adolescents at genetic risk of schizophrenia and bipolar disorder. Prog Neuropsychopharmacol Biol Psychiatry. 2022;112:110432.
24. Morrison EH, Sorkin D, Mosqueda L, Ayutyanont N. Validity and Reliability of the Scale to Report Emotional Stress Signs-Multiple Sclerosis (STRESS-MS) in Assessing Abuse and Neglect of Adults With Multiple Sclerosis. Int J MS Care. 2022;24(1):18-24.
25. Nilsson A, Ibounig T, Lyth J, Alkner B, von Walden F, Fornander L, et al. Protocol: BioFACTS: biomarkers of rhabdomyolysis in the diagnosis of acute compartment syndrome–protocol for a prospective multinational, multicentre study involving patients with tibial fractures. BMJ Open. 2022;12(5)
26. Nyrenius J, Eberhard J, Ghaziuddin M, Gillberg C, Billstedt E. Prevalence of Autism Spectrum Disorders in Adult Outpatient Psychiatry. J Autism Dev Disord. 2022:1-11.
27. Olsson H, Kartasalo K, Mulliqi N, Capuccini M, Ruusuvuori P, Samaratunga H, et al. Estimating diagnostic uncertainty in artificial intelligence assisted pathology using conformal prediction. Nat Commun. 2022;13(1):7761.
28. Ou YC, Tsao TY, Chang MC, Lin YS, Yang WL, Hang JF, et al. Evaluation of an artificial intelligence algorithm for assisting the Paris System in reporting urinary cytology: A pilot study.

Cancer Cytopathol. 2022;130(11):872-80.
29. Paap M, Heltne A, Pedersen G, Germans Selvik S, Frans N, Wilberg T, et al. More is more: Evidence for the incremental value of the SCID-II/SCID-5-PD specific factors over and above a general personality disorder factor. Pers Disord. 2022;13(2):108.
30. Pagnier M, Chaste P. Preliminary Psychometric Properties of the Autism Mental Status Examination (AMSE) in a Tertiary Care Center for Autism Spectrum Disorder Diagnosis. Trends Psychol. 2022:1-22.
31. Papan C, Reifenrath K, Last K, Attarbaschi A, Graf N, Groll AH, et al. Antimicrobial use in pediatric oncology and hematology: protocol for a multicenter point-prevalence study with qualitative expert panel assessment. JMIR Res Protoc. 2022;11(6)
32. Pecukonis M, Young GS, Brian J, Charman T, Chawarska K, Elsabbagh M, et al. Early predictors of language skills at 3 years of age vary based on diagnostic outcome: A baby siblings research consortium study. Autism Res. 2022;15(7):1324-35.
33. Pedersen G, Normann-Eide E, Eikenæs IUM, Kvarstein EH, Wilberg T. Psychometric evaluation of the Norwegian Toronto Alexithymia Scale (TAS-20) in a multisite clinical sample of patients with personality disorders and personality problems. J Clin Psychol. 2022;78(6):1118-36.
34. Peterson BS, Kaur T, Baez MA, Whiteman RC, Sawardekar S, Sanchez-Peña J, et al. Morphological biomarkers in the amygdala and hippocampus of children and adults at high familial risk for depression. Diagnostics. 2022;12(5):1218.
35. Portefaix A, Pons S, Ouziel A, Basmaci R, Rebaud P, Delafay MC, et al. Performance evaluation of host biomarker combinations for the diagnosis of serious bacterial infection in young febrile children: a double-blind, multicentre, observational study. J Clin Med. 2022;11(21):6563.
36. Reiersen AM, Noel JS, Doty T, Sinkre RA, Narayanan A, Hershey T. Psychiatric Diagnoses and Medications in Wolfram Syndrome. Scand J Child Adolesc Psychiatr Psychol. 2022;10(1):163-74.
37. Ridenour JM, Lewis KC, Siefert CJ, Stein MB. Longitudinal stability of Social Cognition and Object Relations Scale—Global Rating Method dimensional ratings, score ranges and narrative 'blandness' in a clinical sample. Clin Psychol Psychother. 2022;29(4):1447-56.
38. Rossoni AM, Lovero KL, Tahan TT, Netto AR, Rossoni MD, Almeida IN, et al. Evaluation of pulmonary tuberculosis diagnostic tests in children and adolescents at a pediatric reference center. Pulmonology. 2022;28(2):83-89.
39. Sadleir PH, Clarke RC, Goddard CE, Mickle P, Platt PR. Agreement of a clinical scoring system with allergic anaphylaxis in suspected perioperative hypersensitivity reactions: prospective validation of a new tool. Br J Anaesth. 2022;129(5):670-8.
40. Sandström J, Myburgh H, Laurent C, Swanepoel DW, Lundberg T. A machine learning approach to screen for otitis media using digital otoscope images labelled by an expert panel. Diagnostics. 2022;12(6):1318.
41. Schrank GM, Sick-Samuels A, Bleasdale SC, Jacob JT, Dantes R, Gokhale RH, et al. Development and evaluation of a structured guide to assess the preventability of hospital-onset bacteremia and fungemia. Infect Control Hosp Epidemiol. 2022;43(10):1326-32.
42. Seidman AJ, George CJ, Kovacs M. Ecological momentary assessment of affect in depression-prone and control samples: Survey compliance and affective yield. J Affect Disord. 2022;311:63-8.
43. Shima C, Lee R, Coccaro EF. Associations of aggression and use of caffeine, alcohol and nicotine in healthy and aggressive individuals. J Psychiatr Res. 2022;146:21-7.
44. Spang KS, Hagstrøm J, Ellersgaard D, Christiani C, Hemager N, Burton BK, et al. Emotion regulation in 7-year-old children with familial high risk for schizophrenia or bipolar disorder compared to controls–The Danish High Risk and Resilience Study–VIA 7, a population-based cohort study. Br J Clin Psychol. 2022;61(4):1103-18.
45. Timmins MA, Berman ME, Coccaro EF. Life history of experienced and witnessed aggression: Development of a new assessment instrument. J Psychiatr Res. 2022;155:518-25.
46. Vieira LS, Nguyen B, Nutley SK, Bertolace L, Ordway A, Simpson H, et al. Self-reporting of psychiatric illness in an online patient registry is a good indicator of the existence of psychiatric

illness. J Psychiatr Res. 2022;151:34-41.
47. Wolff N, Kohls G, Mack JT, Vahid A, Elster EM, Stroth S, et al. A data driven machine learning approach to differentiate between autism spectrum disorder and attention-deficit/hyperactivity disorder based on the best-practice diagnostic instruments for autism. Sci Rep. 2022;12(1):18744.
48. Yang S, Han D, Zhou H, Yang C, Zhang K, Chen S, et al. Validity and Cutoff Score of the Autism Mental Status Exam for an Autism Spectrum Disorder Diagnosis in Chinese Children. J Autism Dev Disord. 2022;1-8.
49. Yang X, Evans RW, George CJ, Matthews KA, Kovacs M. Adiposity and Smoking Mediate the Relationship Between Depression History and Inflammation Among Young Adults. Int J Behav Med. 2022;29(6):787-95.
50. Yano T, Takada T, Fujiishi R, Fujii K, Honjo H, Miyajima M, et al. Usefulness of computed tomography in the diagnosis of acute pyelonephritis in older patients suspected of infection with unknown focus. Acta Radiol. 2022;63(2):268-77.

**Standard reports in 2023 references**

1. Al-Khaled T, Patel SN, Valikodath NG, Jonas KE, Ostmo S, Allozi R, et al. Characterization of errors in retinopathy of prematurity diagnosis by ophthalmologists-in-training in the United States and Canada. J Pediatr Ophthalmol Strabismus. 2023;60(5):337-43.
2. Alqenae FA, Steinke D, Belither H, Robertson P, Bartlett J, Wilkinson J, et al. A Multi-method Exploratory Evaluation of a Service Designed to Improve Medication Safety for Patients with Monitored Dosage Systems Following Hospital Discharge. Drug Saf. 2023;46(10):1021-37.
3. de Best RF, Coppieters MW, van Trijffel E, Compter A, Uyttenboogaart M, Bot JC, et al. Risk assessment of vascular complications following manual therapy and exercise for the cervical region: diagnostic accuracy of the International Federation of Orthopaedic Manipulative Physical Therapists framework (The Go4Safe project). J Physiother. 2023;69(4):260-6.
4. Birkeneder SL, Bullen J, McIntyre N, Zajic MC, Lerro L, Solomon M, et al. The construct validity of the Childhood Joint Attention Rating Scale (C-JARS) in school-aged autistic children. J Autism Dev Disord. 2023:1-17.
5. Dai D, Dong C, Li Z, Xu S. MS-Net: Learning to assess the malignant status of a lung nodule by a radiologist and her peers. J Appl Clin Med Phys. 2023;24(7)
6. Delehanty A, Hooker JL, Wetherby AM. Verbal responsiveness in parents of toddlers with and without autism during a home observation. J Autism Dev Disord. 2023:1-14.
7. Detera-Wadleigh SD, Kassem L, Besancon E, Lopes F, Akula N, Sung H, et al. A resource of induced pluripotent stem cell (iPSC) lines including clinical, genomic, and cellular data from genetically isolated families with mood and psychotic disorders. Transl Psychiatry. 2023;13(1):397.
8. Dimian AF, Estes AM, Dager S, Piven J, Wolff JJ, IBIS Network. Predicting self-injurious behavior at age three among infant siblings of children with autism. Autism Res. 2023;16(9):1670-80.
9. Eldeeb SY, Ludwig NN, Wieckowski AT, Dieckhaus MF, Algur Y, Ryan V, et al. Sex differences in early autism screening using the Modified Checklist for Autism in Toddlers, Revised, with Follow-Up (M-CHAT-R/F). Autism. 2023;27(7):2112-23.
10. Ergül C, Drukker M, Binbay T, Kırlı U, Elbi H, Alptekin K, et al. A 6-year follow-up study in a community-based population: Is neighbourhood-level social capital associated with the risk of emergence and persistence of psychotic experiences and transition to psychotic disorder? Psychol Med. 2023;53(9):3974-86.
11. Gabunia M, Zirakashvili M, Mebonia N, Mikiahvili T, Lomidze G, Leventhal BL, et al. Adaptation of the Strength and Difficulties Questionnaire for Use in the Republic of Georgia. Alpha Psychiatry. 2023;24(4):128.
12. Halabi S, Shiber S, Paz M, Gottlieb TM, Barash E, Navon R, et al. Host test based on tumor necrosis factor-related apoptosis-inducing ligand, interferon gamma-induced protein-10 and C-reactive protein for differentiating bacterial and viral respiratory tract infections in adults: diagnostic accuracy

study. Clin Microbiol Infect. 2023;29(9):1159-65.

13. Harstad E, Hanson E, Brewster SJ, DePillis R, Milliken AL, Aberbach G, et al. Persistence of autism spectrum disorder from early childhood through school age. JAMA Pediatr. 2023;177(11):1197-205.
14. Hill SY, Hostyk J. A whole exome sequencing study to identify rare variants in multiplex families with alcohol use disorder. Front Psychiatry. 2023;14:1216493.
15. Himmelreich JC, Harskamp RE. Diagnostic accuracy of the PMcardio smartphone application for artificial intelligence–based interpretation of electrocardiograms in primary care (AMSTELHEART-1). Cardiovasc Digit Health J. 2023;4(3):80-90.
16. Holingue C, Pfeiffer D, Ludwig NN, Reetzke R, Hong JS, Kalb LG, et al. Prevalence of gastrointestinal symptoms among autistic individuals, with and without co-occurring intellectual disability. Autism Res. 2023;16(8):1609-18.
17. Ikanga J, Patel SS, Roberts BR, Schwinne M, Hickle S, Verberk IM, et al. Association of plasma biomarkers with cognitive function in persons with dementia and cognitively healthy in the Democratic Republic of Congo. Alzheimer's & Dementia: Diagnosis, Assessment & Disease Monitoring. 2023;15(4)
18. Johnson AJ, Shankland E, Richards T, Corrigan N, Shusterman D, Edden R, et al. Relationships between GABA, glutamate, and GABA/glutamate and social and olfactory processing in children with autism spectrum disorder. Psychiatry Res Neuroimaging. 2023;336:111745.
19. Jones W, Klaiman C, Richardson S, Lambha M, Reid M, Hamner T, et al. Development and replication of objective measurements of social visual engagement to aid in early diagnosis and assessment of autism. JAMA Netw Open. 2023;6(9)
20. Kauw F, Velthuis BK, Takx RA, Guglielmo M, Cramer MJ, van Ommen F, et al. Detection of cardioembolic sources with nongated cardiac computed tomography angiography in acute stroke: results from the ENCLOSE study. Stroke. 2023;54(3):821-30.
21. Kocks JW, Cao H, Holzhauer B, Kaplan A, FitzGerald JM, Kostikas K, et al. Diagnostic performance of a machine learning algorithm (Asthma/Chronic Obstructive Pulmonary Disease [COPD] Differentiation Classification) tool versus primary care physicians and pulmonologists in asthma, COPD, and asthma/COPD overlap. J Allergy Clin Immunol Pract. 2023;11(5):1463-74.
22. Kvarstein EH, Frøyhaug M, Pettersen MS, Carlsen S, Ekberg A, Fjermestad-Noll J, et al. Improvement of personality functioning among people treated within personality disorder mental health services: A longitudinal, observational study. Front Psychiatry. 2023;14:1163347.
23. Kvig EI, Nilssen S. Does method matter? Assessing the validity and clinical utility of structured diagnostic interviews among a clinical sample of first-admitted patients with psychosis: A replication study. Front Psychiatry. 2023;14:1076299.
24. Lacroix L, Papis S, Mardegan C, Luterbacher F, L'Huillier A, Sahyoun C, et al. Host biomarkers and combinatorial scores for the detection of serious and invasive bacterial infection in pediatric patients with fever without source. PLoS One. 2023;18(11)
25. Li Z, Guo X, Zhang J, Liu X, Chang R, He M. Using deep leaning models to detect ophthalmic diseases: A comparative study. Front Med (Lausanne). 2023;10:1115032.
26. Makino T, Suzuki F, Nishiyama T, Ishibashi S, Nakamichi H, Iida T, et al. Psychometrics of the kiddie schedule for affective disorders and schizophrenia present and lifetime version for DSM-5 in Japanese outpatients. Int J Methods Psychiatr Res. 2023;32(4)
27. de la Matta M, Alonso-González M, García-Santigosa M, Arance-García M, Sánchez-Peña J, Castro-Liñán LM, et al. Accuracy and comprehensiveness in recording information of a web-based application for preoperative assessment: A prospective observational study. J PeriAnesth Nurs. 2023;38(3):440-7.
28. McNally Keehn R, Enneking B, Ryan T, James C, Tang Q, Blewitt A, et al. Tele-assessment of young children referred for autism spectrum disorder evaluation during COVID-19: Associations among clinical characteristics and diagnostic outcome. Autism. 2023;27(5):1362-76.
29. Moes HR, Ten Kate JM, Portman AT, van Harten B, van Kesteren ME, Mondria T, et al. Timely referral for device-aided therapy in Parkinson's disease. Development of a screening tool.

Parkinsonism Relat Disord. 2023;109:105359.
30. Mooney MA, Neighbor C, Karalunas S, Dieckmann NF, Nikolas M, Nousen E, et al. Prediction of attention-deficit/hyperactivity disorder diagnosis using brief, low-cost clinical measures: a competitive model evaluation. Clin Psychol Sci. 2023;11(3):458-75.
31. Muris P, Büttgens L, Koolen M, Manniën C, Scholtes N, van Dooren-Theunissen W. Symptoms of selective mutism in middle childhood: Psychopathological and temperament correlates in non-clinical and clinically referred 6-to 12-year-old children. Child Psychiatry Hum Dev. 2023:1-12.
32. Nienhuis PH, van Nieuwland M, van Praagh GD, Markusiewicz K, Colin EM, van der Geest KS, et al. Comparing diagnostic performance of short and long [18F]FDG-PET acquisition times in giant cell arteritis. Diagnostics (Basel). 2023;14(1):62.
33. Nylander E, Sparding T, Floros O, Rydén E, Landén M, Hansen S. The quantified behavioural test plus (QbTest+) in adult ADHD. Nord Psychol. 2023;75(1):20-34.
34. Øvstebø RB, Pedersen G, Wilberg T, Røssberg JI, Dahl HSJ, Kvarstein EH. Countertransference in the treatment of patients with personality disorders: A longitudinal study. Psychother Res. 2023:1-15.
35. Paddick SM, Gamassa E, Mwaluwinga N, Lewis G, Duinmaijer A, Urasa S, et al. Preliminary evaluation of a smartphone application (DelApp) for identification of delirium in sub-Saharan Africa. Acta Neuropsychiatr. 2023;1-9.
36. Patel SN, Al-Khaled T, Kang KB, Jonas KE, Ostmo S, Ventura CV, et al. Characterization of Errors in Retinopathy of Prematurity Diagnosis by Ophthalmologists-in-Training in Middle-Income Countries. Journal of Pediatric Ophthalmology & Strabismus. 2023;60(5):344-52.
37. Pedersen G, Kvarstein EH, Wilberg T, Folmo EJ, Burlingame GM, Lorentzen S. The Group Questionnaire (GQ)—Psychometric properties among outpatients with personality disorders. Group Dynamics: Theory, Research, and Practice. 2023;27(2):81.
38. Rios-Olais FA, Gil-Lopez F, Mora-Cañas A, Demichelis-Gómez R. Tumor lysis syndrome is associated with worse outcomes in adult patients with acute lymphoblastic leukemia. Acta Haematologica. 2023;1-11.
39. Rocha Neto HG, Lessa JLM, Koiller LM, Pereira AM, de Souza Gomes BM, Veloso Filho CL, et al. Non-standard diagnostic assessment reliability in psychiatry: a study in a Brazilian outpatient setting using Kappa. European Archives of Psychiatry and Clinical Neuroscience. 2023;1-12.
40. Schamberg G, Calder S, Varghese C, Xu W, Wang WJ, Ho V, et al. Comparison of Gastric Alimetry® body surface gastric mapping versus electrogastrography spectral analysis. Sci Rep. 2023;13(1):14987.
41. Schulte-Rüther M, Kulvicius T, Stroth S, Wolff N, Roessner V, Marschik PB, et al. Using machine learning to improve diagnostic assessment of ASD in the light of specific differential and co-occurring diagnoses. Journal of Child Psychology and Psychiatry. 2023;64(1):16-26.
42. Smit MR, Hagens LA, Heijnen NF, Pisani L, Cherpanath TG, Dongelmans DA, et al. Lung ultrasound prediction model for acute respiratory distress syndrome: a multicenter prospective observational study. Am J Respir Crit Care Med. 2023;207(12):1591-601.
43. Snelling PJ, Jones P, Bade D, Bindra R, Byrnes J, Davison M, et al. Ultrasonography or radiography for suspected pediatric distal forearm fractures. New England Journal of Medicine. 2023;388(22):2049-57.
44. Spangenberg H, Ramklint M, Ramirez A. A long-term follow-up study of labor market marginalization in psychiatric patients with and without personality disorder. Upsala Journal of Medical Sciences. 2023;128.
45. Susko M, Armstrong VL, Brian JA, Bryson SE, Kushki A, Sacrey LAR, et al. Behavioural reactions to an emotion evoking task in infants at increased likelihood for autism spectrum disorder. Infant Behavior and Development. 2023;72:101848.
46. Sveen CA, Pedersen G, Hummelen B, Kvarstein EH. Personality disorders: the impact of severity on societal costs. European Archives of Psychiatry and Clinical Neuroscience. 2023;1-12.
47. Sveen CA, Pedersen G, Ulvestad DA, Zahl KE, Wilberg T, Kvarstein EH. Societal costs of

personality disorders: A cross-sectional multicenter study of treatment-seeking patients in mental health services in Norway. Journal of Clinical Psychology. 2023;79(8):1752-69.

48. Tärnhäll A, Björk J, Wallinius M, Gustafsson P, Billstedt E, Hofvander B. Healthcare utilization and psychiatric morbidity in violent offenders: findings from a prospective cohort study. Social Psychiatry and Psychiatric Epidemiology. 2023;58(4):617-28.
49. Tärnhäll A, Björk J, Wallinius M, Gustafsson P, Hofvander B. Offending trajectories in violent offenders: criminal history and early life risk factors. International Journal of Offender Therapy and Comparative Criminology. 2023;67(2-3):270-90.
50. Wilberg T, Pedersen G, Bremer K, Johansen MS, Kvarstein EH. Combined group and individual therapy for patients with avoidant personality disorder—a pilot study. Frontiers in Psychiatry. 2023;14:1181686.
51. Yang S, Han D, Zhou H, Yang C, Zhang K, Chen S, et al. Validity and Cutoff Score of the Autism Mental Status Exam for an Autism Spectrum Disorder Diagnosis in Chinese Children. Journal of Autism and Developmental Disorders. 2023;53(12):4822-9.
